# NeMMo: an improved statistical algorithm for excess all-cause mortality surveillance and monitoring

**DOI:** 10.64898/2026.08.14.26360477

**Authors:** Theodore Lytras, Maria Athanasiadou

## Abstract

**Background:** Reliable estimation of excess mortality is central to population health surveillance. We introduce NeMMo (New Mortality Model), an evolution of the EuroMOMO model for estimating weekly all-cause expected mortality, and assess its behaviour and performance on empirical data.

**Methods:** NeMMo incorporates population offsets, stratifies observed deaths by age group and models seasonality using a periodic B-spline rather than a Serfling-type sinusoidal function. Baseline weeks are selected by a data-driven procedure minimizing the skewness of the residuals before refitting the model, instead of relying solely on fixed calendar windows. NeMMo enables pooling across age groups, direct age standardization and incorporation of external predictors. We applied NeMMo and EuroMOMO to mortality and population data downloaded from Eurostat for 31 countries from 2015 onwards, excluding the COVID-19 pandemic period from baseline estimation.

**Results:** For most countries NeMMo produced a higher expected mortality baseline that better tracked observed deaths, as well as tighter prediction intervals and higher maximum Z-scores, suggesting improved discrimination of mortality excesses. Z-scores and P-scores during non-pandemic weeks were closer to zero with NeMMo than with EuroMOMO but further elevated during pandemic weeks, providing greater separation between pandemic and non-pandemic mortality. Incorporating population offsets resulted in negative linear trends across all countries, consistent with declining mortality after accounting for demographic changes. The periodic B-spline identified substantial heterogeneity in the shape and timing of seasonal mortality that was not captured by a sinusoidal function.

**Conclusions:** NeMMo provides a flexible and parsimonious framework for all-cause mortality surveillance that improves the established EuroMOMO model and offers theoretical, empirical and practical advantages. It is thus suitable both for detecting short-term spikes and for the long-term, age-adjusted quantification and comparison of mortality excesses that has become increasingly important since the COVID-19 pandemic. The accompanying ‘nemmo’ package for R facilitates its widespread adoption and application.

## Background

Reliable estimation of excess mortality is a cornerstone of public health surveillance, providing crucial, real-time insights into the burden of health crises such as pandemics, extreme weather events, and seasonal influenza [1,2]. For almost two decades, the European Mortality Monitoring (EuroMOMO) network and its standardized algorithm have successfully served as a beacon for tracking these trends across Europe [3,4]. Following the COVID-19 pandemic, which brought the challenges of excess mortality estimation into the spotlight, it is time for a rethink of not just algorithms but the core concept: what excess mortality is, what we may use it for, and how to best meet those objectives.

Excess mortality has been defined as the difference between observed and expected deaths under some counterfactual [5]. The problem is estimating this fundamentally unknowable counterfactual, and different methods for this can produce divergent excess mortality estimates [6]. For example, EuroMOMO and CDC model expected deaths under a Serfling model of an annual sinusoidal term on top of a linear trend [3,7], whereas Eurostat uses the average monthly deaths over the preceding three years [8]. Both choices have weaknesses; factors like demographic ageing mean future deaths are unlikely to be equal to past deaths, nor is there an inherent reason for baseline mortality to follow a sinusoidal pattern. Conceptually the target of estimation for this counterfactual of future expected deaths can be one of two things, that directly influence its interpretation: (a) a “natural” level of mortality that excludes any deaths deemed “unnatural”, or (b) a “usual”, expected level that will inevitably incorporate some regular and recurrent influences on mortality such as respiratory virus activity. The former is probably unattainable without cause-of-death data, not to mention the elusiveness of defining what a “natural” death is. We are therefore left with the second paradigm: defining the counterfactual by isolating a predictable historical pattern and extrapolating as best as possible into the future.

The next question is what Public Health objectives excess mortality estimates are used for. The original purpose of EuroMOMO was detection of short-term mortality spikes, which may correspond to events like a unusually intense influenza season or a heatwave or the start of a pandemic [3]. But COVID-19 highlighted the need for reliable estimation of long-term deviations too, for the purpose of quantifying the mortality burden of the pandemic and making comparisons between countries, age groups and different periods [4]. This places a premium on age-adjustment, as an essential requirement for valid comparisons but also for distinguishing true long-term mortality excesses from structural shifts in baseline mortality, which a linear trend cannot sufficiently capture. In this context, including population sizes improves calculation of expected mortality and facilitates both age-adjustment and age-stratification of mortality excesses, which is important for Public Health decisionmaking. In addition, including population size in a model for expected mortality “frees” its linear trend to indicate true long-term change in underlying mortality rates rather than demographic changes. The periodic terms of the model are interesting too, especially compared between age groups, in revealing the magnitude and timing of the seasonal variation in mortality; for this purpose, modern approaches such as splines may offer a better balance between accuracy, parsimony and interpretability compared to traditional sinusoidal terms [9,10].

The recent emphasis on longer-term mortality trends has also led to the introduction of P-scores as an alternative measure to the Z-score, arguing for easier interpretability and comparability of excess mortality statistics [11]. In reality both measures are needed: just like effect sizes and p-values are complementary, P-scores express the relative size of a mortality excess while Z-scores express how unusual or unexpected a given excess is. Neither measure obviates the need for appropriate age-adjustment [12], and neither is sufficient on its own to make comparisons and rankings, an endeavour that carries its own complexities [13]. In practice Z-scores are more suitable as short-term alert signals and P-scores for quantifying mortality excesses, making them complementary.

Consequently, and using the EuroMOMO algorithm as a starting point, this paper and accompanying R package proposes an improved algorithm for estimating excess mortality in the context of European all-cause mortality surveillance. We then proceed to apply this new algorithm (called “NeMMo” – New Mortality Model) in analyzing available mortality data from Eurostat, and compare the obtained results against the EuroMOMO model.

## Methods

### Description of the NeMMo model

Similar to the EuroMOMO model, the NeMMo model is a quasi-Poisson model of weekly counts of deaths *Y_kt_*, with log link and a linear term *t* for trend. However, it is by default age-stratified, it includes population size *N_kt_* by default as an offset, and replaces the Serfling-type sinusoidal terms with a periodic B-spline function of week number B(*w_t_*) with three internal knots. Thus for every age group *k* we have:

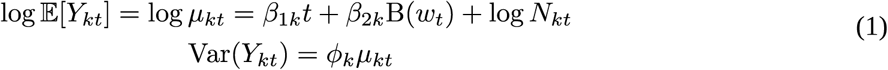

where *ϕ_k_*is the overdispersion parameter, and *β_2k_* becomes a parameter vector of length three.

In Europe, population size of age group *k* is usually known or projected by national statistical services for the first week of each year; we propose using the latest three available values to linearly regress as needed into the future, and to interpolate the values for all weeks in-between using a cubic spline with knots at every known value (i.e. the start of each year). This is a reasonable way to estimate *N_kt_* for the entire series, thereby freeing the *β_1k_* parameter from capturing demographic trends. In addition, a correction for reporting delay can be applied to the counts of observed deaths *Y_kt_* if desired, based on the proportion *r_d_* of deaths that are historically reported within *d* weeks: *Y_kt_* = *Y^0^* / *r_d_* where *t* = *T* – *d*. If performed, *r_d_* should be estimated separately using any predictors deemed relevant.

The observed counts *Y_kt_* will undoubtedly include periods of excess mortality due to influenza, heatwaves or other unusual effects; these need to be trimmed out in order to estimate a true mortality baseline *μ_kt_*. The EuroMOMO algorithm handles this problem by including only spring and autumn weeks in its model (ISO weeks 16-25 and 37-44 respectively) [14]; therefore the other weeks do not contribute at all to the estimation of excess mortality, and their baseline is being extrapolated using the sinusoidal term. NeMMo introduces an alternative approach: first fit the model of equation (1) to the entire time series of weekly observed deaths and obtain the model deviance residuals *e_t_*, which will be right-skewed as a result. Then start trimming the highest residuals in order to identify the set of weeks *T̂* that minimizes the skewness of the residuals, i.e. the difference between their mean and median, making them as symmetric as possible:

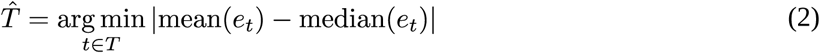

Then refit equation (1) using only *t* ∈ *T̂*, i.e. those weeks that create a truly symmetric expected mortality baseline, with upward deviations just as likely as downward deviations. The initial set *T* can and should also be manually trimmed if there is any a priori known atypical period, such as a pandemic, which should not be used to inform an expected mortality level. But in addition to that, this single-step refitting approach immediately filters out the remaining excesses to create a meaningful baseline, while utilizing more of the available data and better informing the periodic B-spline component of the baseline.

The above “symmetrizing the residuals” approach has a limitation for sparse data, especially when there are many zero-count observations. In such cases it “filters out” weeks with non-zero deaths leaving an excessively low baseline (often zero). Therefore, for groups with a mean weekly count of E(λ)<5 we apply a more conservative filter: trim only those weeks with an observed count higher than the 99%-th percentile of the corresponding Poisson distribution (e.g. for a mean of 4, higher than 9) and then refit. Alternatively one may choose to merge groups into broader categories to increase counts, though that will weaken adjustment and introduce residual confounding.

For every group *k*, the expected mortality baseline is formed by the model-predicted deaths *μ_kt_*; their standard errors in the log scale SE(log *μ_kt_*) and the overdispersion parameter *ϕ* are also estimated by the model. To calculate a Z-score for the observed deaths *Y_kt_* the same 2/3-power transformation is applied as in EuroMOMO, therefore:

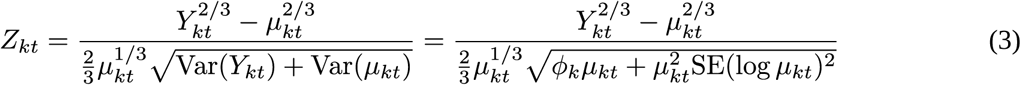

and to calculate an upper prediction limit *U* that should be *L*>0 standard deviations from the baseline, we have:

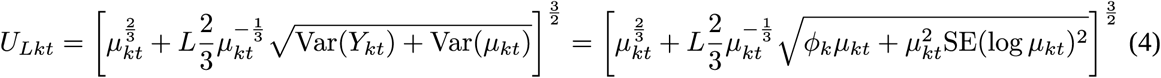

Interestingly, given independence between groups (since they pertain to different people), equations (3) and (4) apply also for the sum of observed deaths across all groups *K*. All we have to do is sum together the observed and expected counts, as well as the process variance Var(*Y_kt_*) and estimation variance Var(*μ_kt_*):

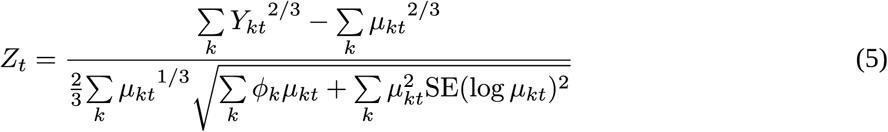

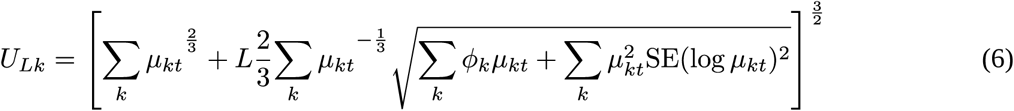

There is therefore no need for a model fit on total deaths in a population; both baseline and prediction intervals can be calculated by pooling estimates from group fits. In addition, it is possible to pool together the parameters from the group fits (the linear trend *β_1k_* or the periodic B-spline *β_2k_*) weighted by any desired weight *w_k_*, for example the share of group-specific deaths among total deaths in the time series:

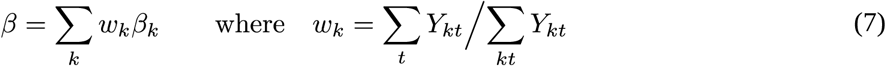

This allows studying mortality trends and seasonal patterns both overall and comparatively between age groups. It also allows direct age adjustment by applying population weights (equal to the ratio between reference population and study population) to the observed deaths, expected deaths, process and estimation variance, before summing up everything together as above:

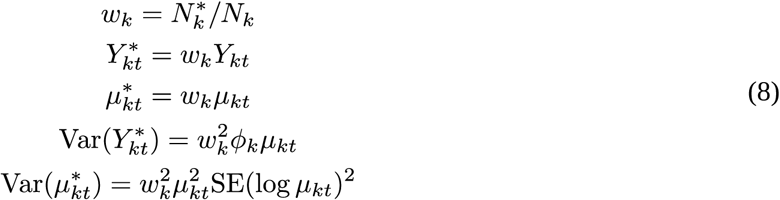

where *N \**is the size of the reference population subgroup *k*. In this way observed, expected and excess mortality can be compared between different countries if desired, as well as within a country over time eliminating the effect of long-term shifts in its age structure.

Finally, the model can incorporate an additional matrix of predictors P, which is useful to examine and quantify external influences on mortality, such as temperature, respiratory virus activity and others. Therefore the same model can be used to detect short-term extremes, monitor longer-term deviations and estimate attributable mortality, all while taking account of demographic shifts.

### Analysis of all-cause mortality in Europe with the NeMMo model

To examine the behaviour and performance of the NeMMo model, we applied it side-by-side with the EuroMOMO model on death and population data for 31 European countries downloaded from Eurostat (in August 2026), starting from the first week of 2015. We trimmed the last four available weeks for each country to minimize reporting delay concerns, and manually excluded the pandemic period between week 1/2020 and 20/2023 from baseline calculation. For the NeMMo model we applied age-stratification in 5-year groups (from 0-4 up to 85+ years) for both sexes together, whereas EuroMOMO was applied to total deaths without age stratification.

We visually examined the generated baseline, prediction intervals, Z-scores and P-scores for all countries, and compared the distribution of Z-scores and P-scores, the level of the baseline and the amplitude of the prediction intervals, between NeMMo and EuroMOMO. We further compared the mean (rather than cumulative) P-scores and Z-scores between pandemic weeks (1/2020 to 20/2023) and non-pandemic weeks for both the NeMMo and EuroMOMO models, in order to assess how clearly each model distinguishes the impact of the COVID-19 pandemic. Finally, we compared the effect of the linear trend between both models, and examined the more flexible seasonality predicted by the NeMMo model for each country. All analyses were done in R [15], with full code available at https://github.com/thlytras/analysis_nemmo_eurostat.

### Implementing the NeMMo model in R: the nemmo package

To facilitate our analysis and the implementation of the NeMMo model by others for research and mortality surveillance purposes, we created the ‘nemmo’ package for the R statistical environment. This is available on https://github.com/thlytras/nemmo and on the Comprehensive R Archive Network (CRAN). The package is centered around objects of class ‘mmort’ (mortality model) which can fit models of both NeMMo and EuroMOMO specification. These models are essentially R lists that hold all associated data, and have methods for plotting, calculating baselines and prediction intervals, extracting effects (linear, seasonality and predictor effects), subsetting (e.g. to select particular age groups in a fully age-stratified NeMMo model), pooling (to pool together age-groups within a model and across different models fitted to the same period) and for direct age adjustment. By abstracting these operations, the ‘mmort’ class and its methods greatly simplify writing R code for surveillance and analysis purposes. The ‘nemmo’ package also includes (as associated data) all the deaths and population data from Eurostat that we used for analysis, as well as the European Standard Population 2013 which is a sensible choice for direct age adjustment of ‘mmort’ class objects. The package also includes comprehensive documentation that explains its use.

## Results

The characteristics of the 31 analyzed countries (with available complete deaths and population data from Eurostat) are listed in Table 1, encompassing a diverse variety of countries large and small; weekly population was interpolated from annual population data as described above. NeMMo and EuroMOMO model fits from each country, in terms of expected mortality, Z-scores and P-scores, are illustrated in Supplementary Figure 1 (an indicative example from Cyprus in Figure 1) with comparative summaries in Table 2.

**Figure 1:**
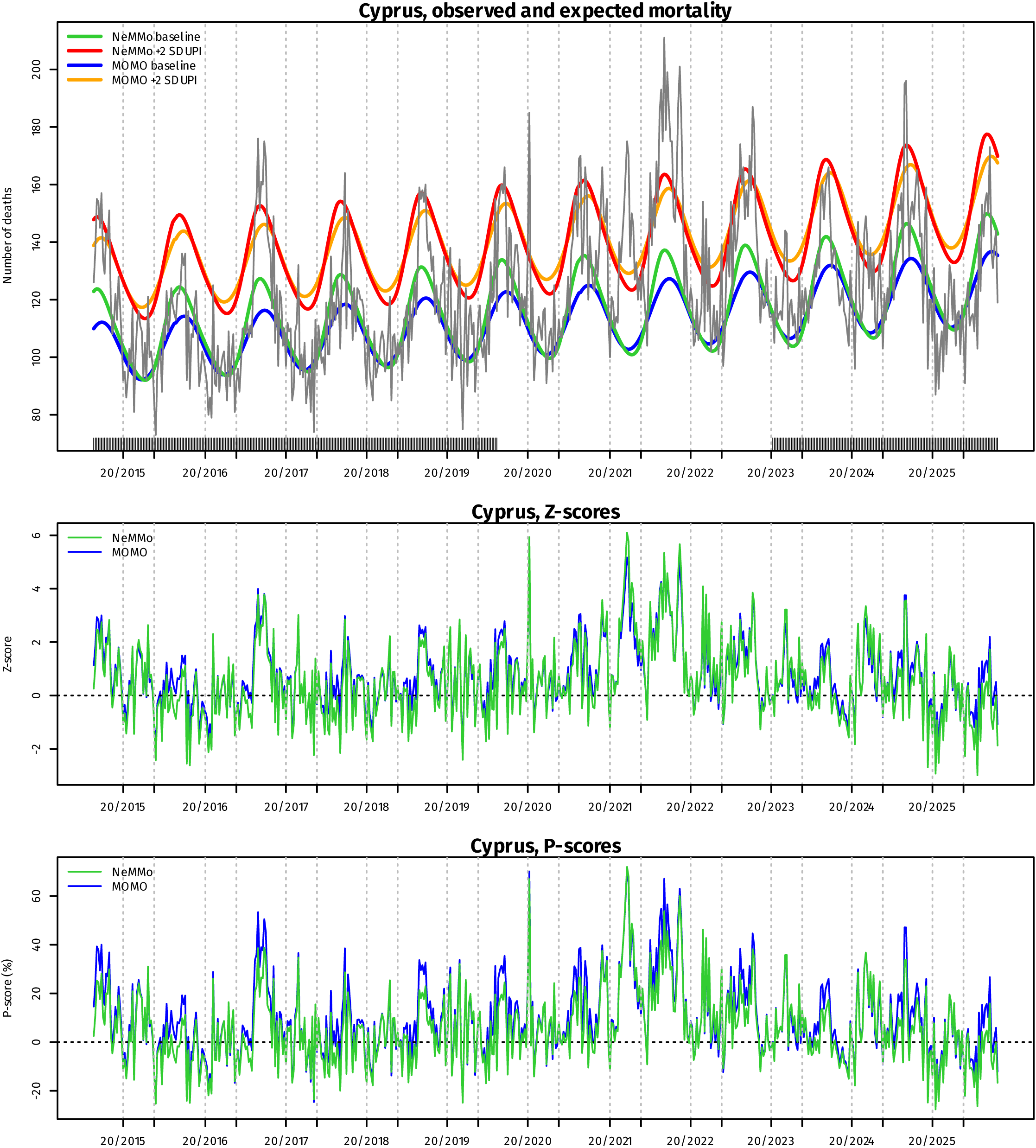
**Observed and expected mortality, Z-scores and P-scores between NeMMo and EuroMOMO for Cyprus (for all countries, see Supplementary Figure 1)**

**Table 1:** Descriptive characteristics of analyzed countries.

| Country | Weekly population | Weekly deaths (all ages) |
| --- | --- | --- |
| Austria | 8,925,025 (8,584,926–9,263,947) | 1592 (1285–2544) |
| Belgium | 11,546,061 (11,237,274–11,983,314) | 2082 (1682–4292) |
| Bulgaria | 6,541,467 (6,429,432–7,029,690) | 2023 (1604–4753) |
| Croatia | 3,910,166 (3,850,839–4,180,915) | 989 (781–1785) |
| Cyprus | 909,197 (860,846–1,002,664) | 119 (73–211) |
| Czechia | 10,627,077 (10,455,688–10,955,289) | 2134 (1761–4238) |
| Denmark | 5,833,937 (5,659,715–6,035,216) | 1059 (879–1429) |
| Estonia | 1,328,891 (1,313,271–1,375,050) | 304 (230–438) |
| Finland | 5,529,834 (5,471,753–5,679,503) | 1078 (875–1483) |
| France | 67,634,406 (66,458,153–69,305,182) | 11,756 (9926–18,802) |
| Greece | 10,593,049 (10,361,848–10,818,041) | 2368 (1849–3895) |
| Hungary | 9,669,698 (9,500,441–9,815,858) | 2454 (1935–4568) |
| Iceland | 366,368 (329,100–392,101) | 45 (25–78) |
| Italy | 59,410,648 (58,912,366–60,295,497) | 12,356 (10,321–23,457) |
| Latvia | 1,900,895 (1,841,042–1,986,096) | 538 (424–927) |
| Liechtenstein | 38,978 (37,366–41,772) | 5 (0–25) |
| Lithuania | 2,852,312 (2,802,677–2,926,644) | 760 (555–1395) |
| Luxembourg | 631,283 (562,958–694,238) | 83 (49–137) |
| Malta | 516,093 (438,805–592,606) | 73 (37–133) |
| Montenegro | 622,200 (616,036–626,247) | 124 (85–261) |
| Netherlands | 17,449,516 (16,900,726–18,175,774) | 3044 (2492–5085) |
| Norway | 5,379,678 (5,165,802–5,640,192) | 802 (662–1137) |
| Poland | 37,423,588 (36,340,995–38,051,722) | 7770 (6554–16,244) |
| Portugal | 10,391,151 (10,329,955–10,883,631) | 2138 (1697–5039) |
| Romania | 19,281,402 (19,031,726–19,870,647) | 4892 (3884–11,641) |
| Serbia | 6,891,507 (6,514,635–7,114,393) | 1903 (251–4088) |
| Slovakia | 5,431,612 (5,413,978–5,462,228) | 1024 (855–2125) |
| Slovenia | 2,105,062 (2,062,874–2,139,042) | 395 (303–801) |
| Spain | 47,393,484 (46,413,752–49,865,778) | 8065 (5462–20,967) |
| Sweden | 10,358,335 (9,747,355–10,626,895) | 1686 (1429–2569) |
| Switzerland | 8,644,512 (8,237,666–9,190,655) | 1306 (1078–2111) |

From the original Eurostat dataset (demo\_r\_mwk2\_05), the following countries were excluded: Germany (no age-specific data below age 40), Ireland (no age-specific data at all), the United Kingdom (data stop at the end of 2020) and Albania (data stop at week 37/2021)

**Table 2:**
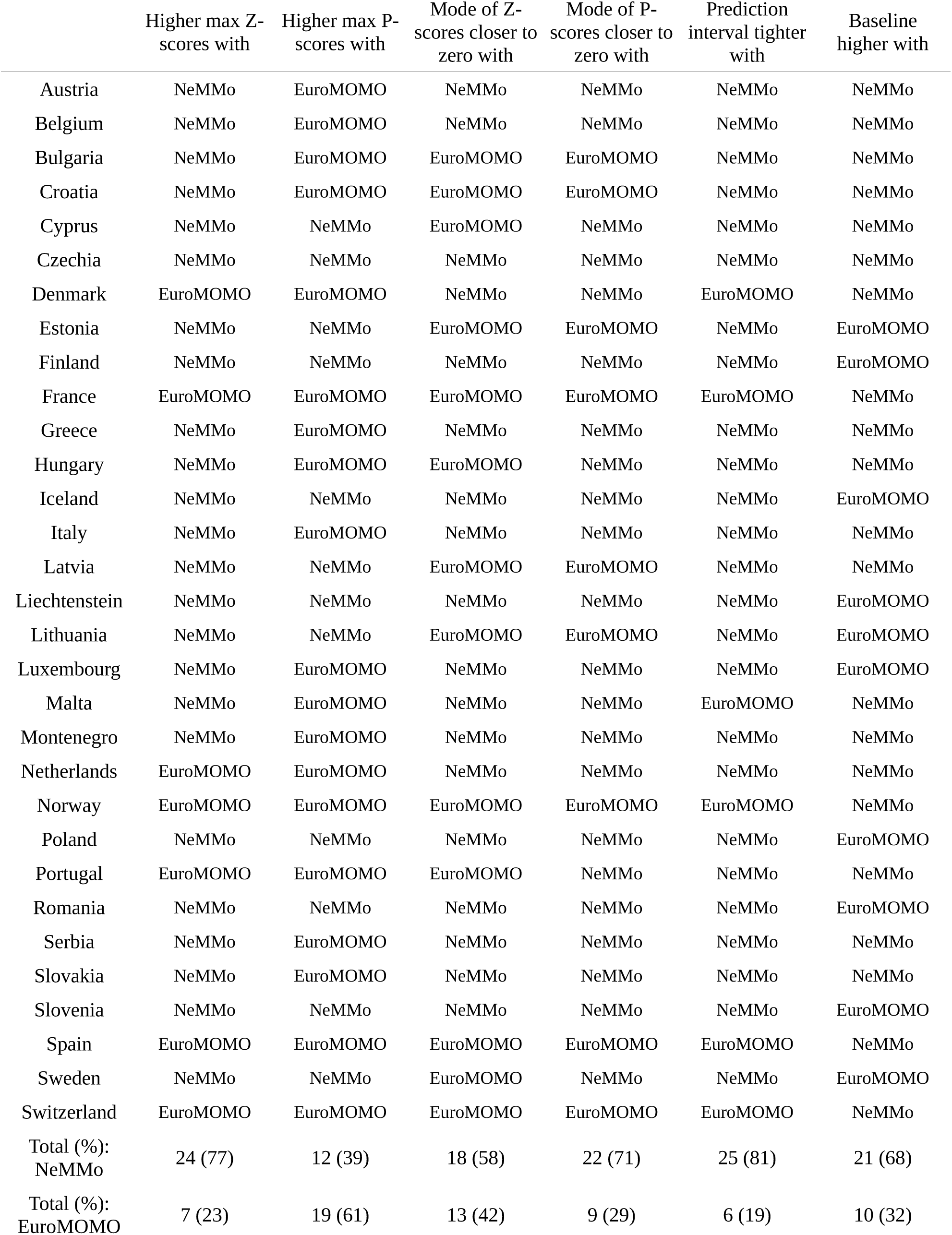
Comparative summary of NeMMo and EuroMOMO model fits.

Although results are largely similar, the NeMMo expected mortality baseline visually appears to track off-season observed deaths closer than EuroMOMO in most cases, and it was on average higher than the EuroMOMO baseline in 21 countries (68%) and lower in 10 (32%). Importantly, 24 countries (77%) had higher maximum Z-scores with NeMMo than with EuroMOMO, mostly due to tighter prediction intervals (25 countries, Figure 2) resulting in better effective discrimination of mortality excesses. Countries having higher maximum Z-scores in EuroMOMO instead of NeMMO (Denmark, France, Netherlands, Norway, Portugal, Spain and Switzerland) only did so as a result of a lower baseline in EuroMOMO compared to NeMMo (Table 2). At the same time the mode of the Z-score distribution with the NeMMo model was closer to zero for the majority of countries (18/31), suggesting a more symmetric distribution; the same was not true of P-scores, primarily as a result of a higher NeMMo baseline. A full comparison between NeMMo and EuroMOMO in terms of the Z-score and P-score distributions for every country is provided in Supplementary Figure 2.

**Figure 2:**
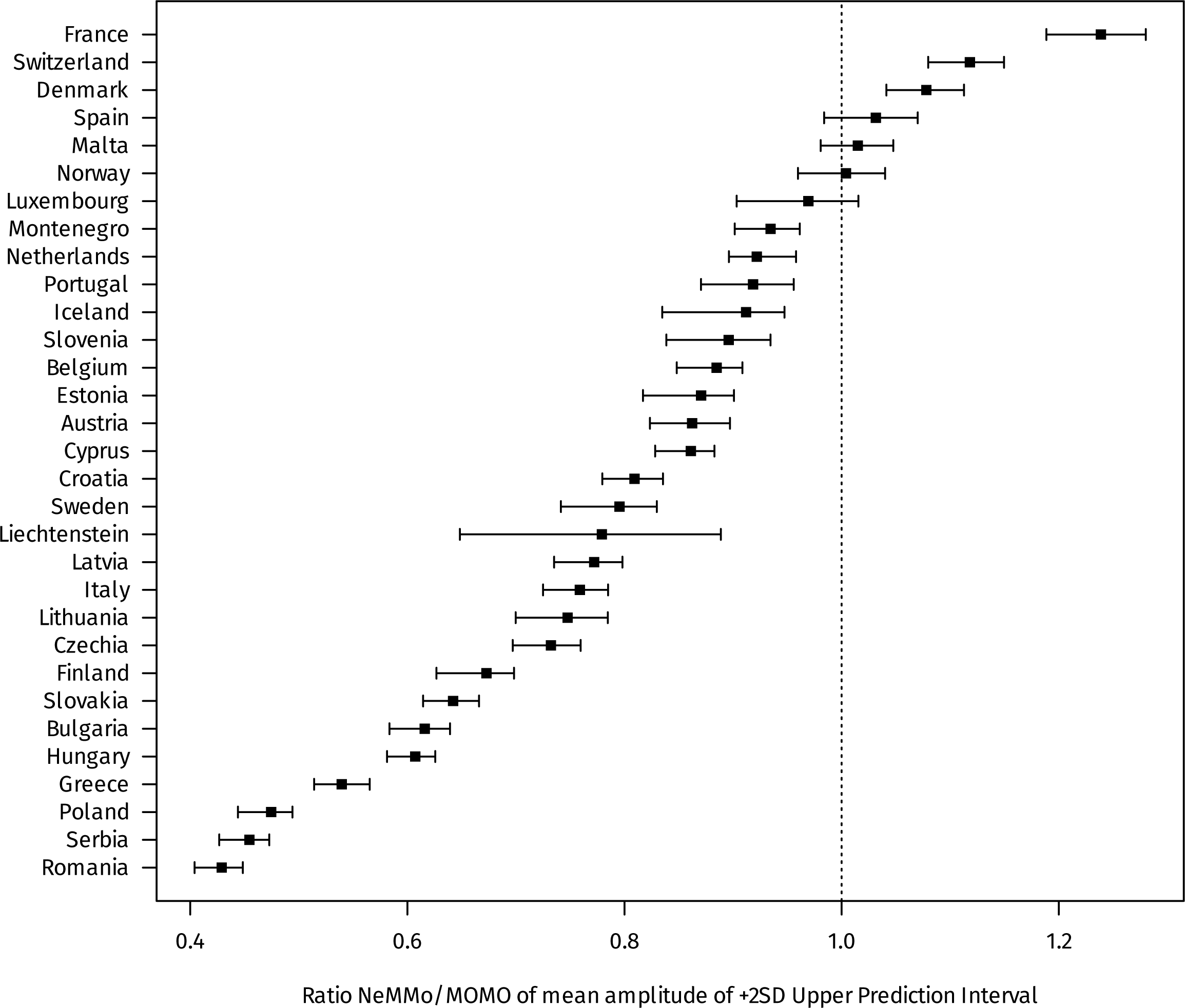
**Amplitude ratio of +2SD Upper Prediction Interval between NeMMo and EuroMOMO, all countries**

We then examined the performance of the NeMMo and EuroMOMO algorithms in the context of the COVID-19 pandemic, by calculating the mean weekly P-scores and Z-scores during the pandemic weeks (01/2020 to 20/2023) and comparing with the non-pandemic weeks (Figure 3). For most countries the non-pandemic P-scores are closer to zero with NeMMo than EuroMOMO (mean +1.6% vs +3.0%), allowing a clearer separation between non-pandemic and pandemic weeks. The same occurs with Z-scores, with a mean non-pandemic Z-score of +0.36 with NeMMO and +0.55 for EuroMOMO, and mean pandemic scores of +2.94 and +2.29 respectively, primarily as a result of the tighter prediction intervals with NeMMo. Therefore the NeMMo algorithm appears to discriminate true excess mortality slightly but meaningfully better than the EuroMOMO algorithm.

**Figure 3:**
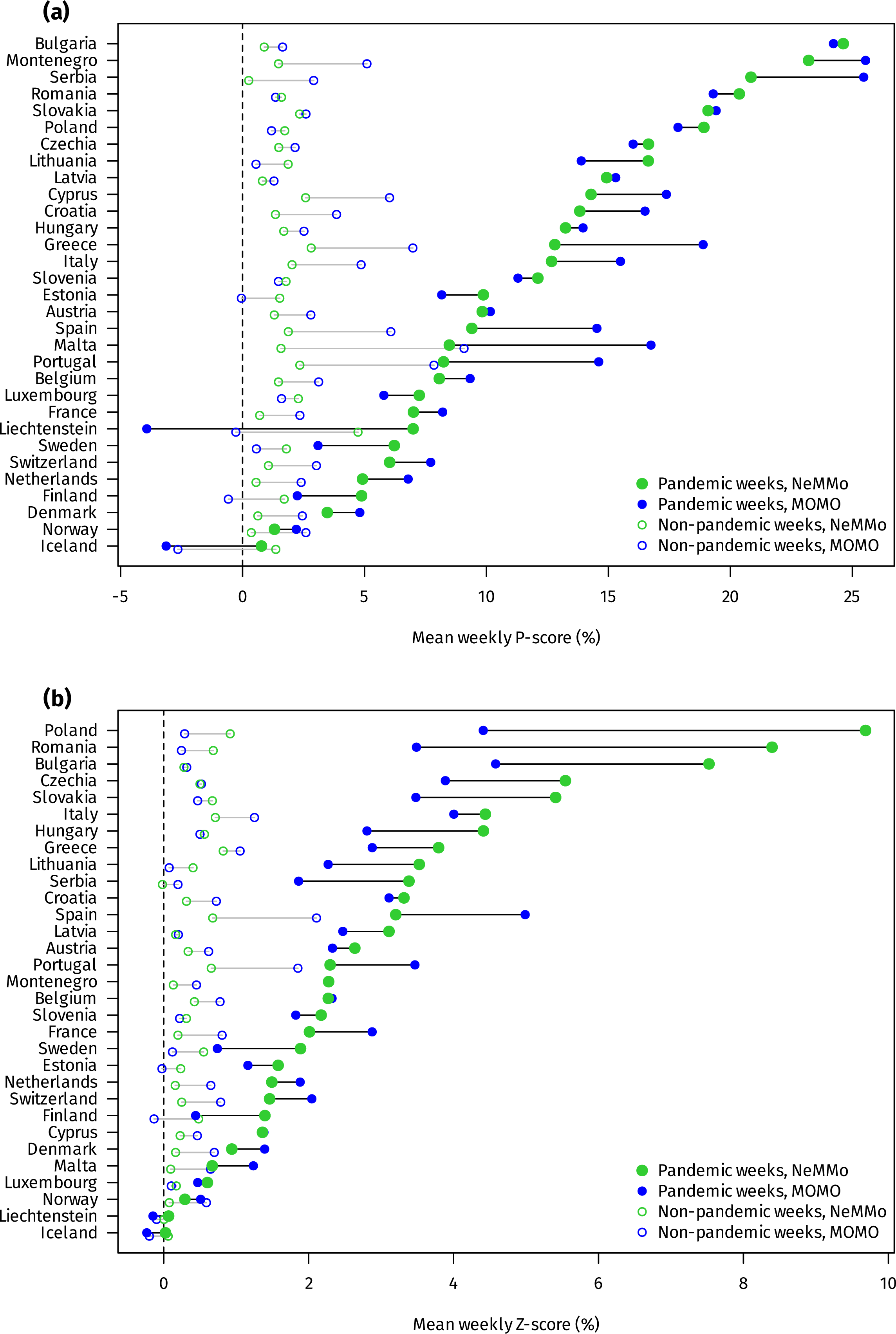
**Mean weekly P-scores and Z-scores for pandemic and non-pandemic weeks across all countries, NeMMo vs EuroMOMO**

There are other advantages with NeMMo as well. Figure 4 illustrates *β_1_*, the effect of the linear trend for both NeMMo (pooled across all age groups) and EuroMOMO across the studied countries. With EuroMOMO most betas are positive, especially for countries with growing and ageing populations. With NeMMo on the other hand, and as a result of the included population offset, all betas are negative reflecting a true improvement in overall mortality rates over time. Therefore *β_1_* with NeMMo is interpretable as the true long-term effect on expected mortality (conditional on correct estimation of population sizes), whereas *β_1_* with EuroMOMO is confounded by demographic trends and cannot be interpreted as such. Figure 5 similarly illustrates the seasonality effect on mortality as modelled with NeMMo by country. For some countries the effect is close to sinusoidal, but for many it is not (such as Austria, France, Norway, Serbia, Switzerland and others), with a larger mortality peak in winter and a much smaller trough in summer. The timing of the winter peaks and summer troughs also varied by country, and for the most part were not equidistant as implied by the sinusoidal curve. Thus the periodic B-spline employed by the NeMMo model offers a more flexible and realistic modelling of the seasonal part of expected mortality, while still being parsimonious (with only three regression coefficients).

**Figure 4:**
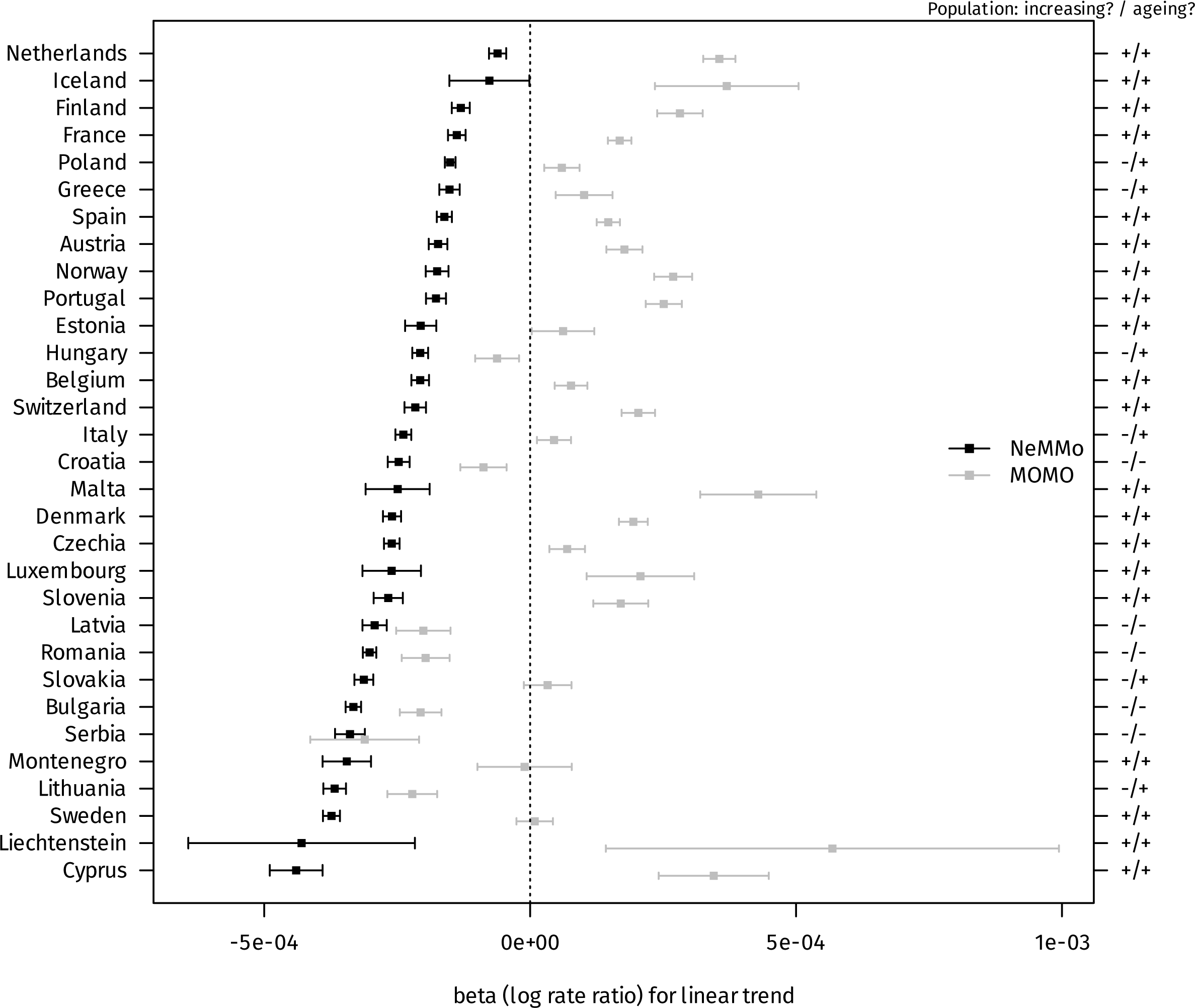
**Effect of linear trend (mean and 95% Confidence Interval) on expected mortality, NeMMo vs EuroMOMO, all countries**

**Figure 5:**
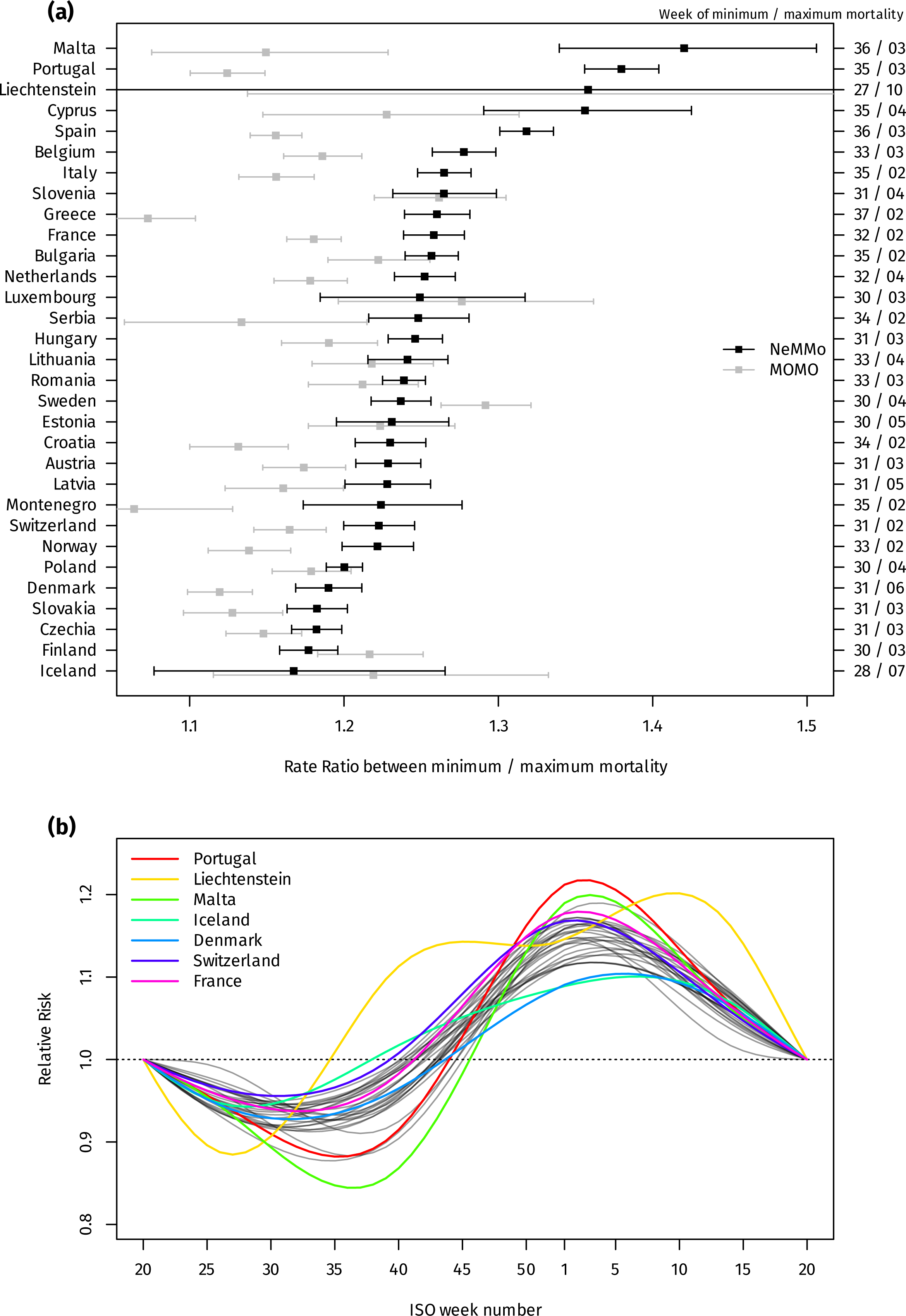
**Seasonality effect on expected mortality, all countries. (a) Peak-to-trough Rate Ratio, NeMMo vs EuroMOMO. (b) Mortality Rate Ratios by week (centered at week 20), NeMMo model, all countries.**

## Discussion

In this paper we introduce NeMMo, an evolution of the EuroMOMO algorithm for surveillance of all-cause mortality, and compare it against the original EuroMOMO using country data downloaded from Eurostat. Four major changes distinguish NeMMo from its predecessor: the “symmetrizing of residuals” procedure to filter excess weeks, the switch to a periodic B-spline instead of a Serfling-type sinusoidal curve, the default inclusion of population offsets and group stratification (primarily age group stratification, though this can be applied to any type of group). These changes result in substantial theoretical, empirical and practical advantages.

On a theoretical level, the residual-symmetrizing procedure creates a baseline with a coherent interpretation: for non-epidemic weeks, mortality is as likely to be above the baseline as it is to be below it. The procedure is data-driven, utilizing much more of the time-series than the fixed-window of EuroMOMO (weeks 16-25 and 37-44). The periodic B-spline allows a more flexible modelling of seasonality, including the shape and timing of the winter peaks and summer troughs, rather than extrapolating those through a weakly anchored sinusoidal curve. The result is a conceptually more defensible baseline, that empirically appears to better track baseline mortality off-season in our data, while still better distinguishing true mortality excesses both during short-term spikes and during the long COVID-19 pandemic period.

The robustness of the baseline is further enhanced by the inclusion of population offsets. For short-term spike detection this may not be so significant, as demographic shifts are largely captured by the linear trend. But over longer periods of time (such as during a pandemic) changes in population size and age structure can meaningfully bias expected mortality upwards or downwards, and thus should be taken into account. Including population offsets has the additional benefit of revealing true long-term mortality trends by disentangling the effect of the linear trend from population dynamics, as was apparent in our data. The same informedness was true of the flexible B-spline modelling of seasonality, revealing a pattern of larger mortality peaks in winter and shallower mortality troughs in summer which has been observed before [10], and might become more relevant in the future given the health effects of global warming [16]. The B-spline also reveals the timing of the winter peak and summer trough, which is variable in different countries and not captured by the sinusoidal trend. Therefore the NeMMo model offers valuable additional insights that are useful for Public Health decisionmaking.

The use of age stratification further improves the reliability of the pooled baseline and produces tighter prediction intervals, as was evident in our analysis, by limiting overdispersion in the quasi-Poisson model fits [17]. It also offers the ability to pin any short- or long-term mortality excesses to particular age groups, and to perform direct age adjustment if needed. Finally the optional inclusion of additional predictors in the NeMMo model provides a practical avenue to study mortality attributable to external influences like respiratory viruses and temperature. This is significant because these exposures are not only associated with “unexpected” excess deaths, but account for part of the expected seasonal variation in mortality [18]; therefore using excess deaths estimates as outcome in modelling attributable mortality only looks at the tip of the iceberg, and it is important to provide a practical unified framework to study both at the same time, which is what the NeMMo model and our ‘nemmo’ R package do. This will be an avenue for future research.

Our work has limitations. We cannot validate the NeMMo model (or EuroMOMO) against a true gold standard because such a standard for expected or excess deaths does not exist [19]; the different approaches to estimating expected mortality all rely on different assumptions [9], and NeMMo only offers a set of simple yet principled and intuitive assumptions (based on the idea of symmetry of the residuals). Our comparison between pandemic and non-pandemic periods did show better discrimination with NeMMo in terms of both P-scores and Z-scores, yet it is still an imperfect test of performance given the presence of weeks with and without excess deaths in both periods. We did not touch upon the issue of reporting delay correction (and only excluded the most recent weeks), which is very relevant for real-time surveillance of short-term mortality spikes; the NeMMo model is open to the implementation of various plug-in corrections, with correct propagation of their uncertainty in Z-score estimation (and this can be implemented in a future version of the ‘nemmo’ R package). Finally, our work is only relevant for countries with sufficient civil registration and vital statistics systems, which should not be a problem for Europe but limits the applicability of the NeMMo model to less developed settings.

In conclusion, our proposed NeMMo algorithm is a meaningful improvement over EuroMOMO with distinct advantages, that performs well in a diverse set of countries analyzed (ranging in size from Liechtenstein to France) and is appropriate not only for short-term mortality surveillance but also for the long-term, age-adjusted quantification and comparisons that excess mortality surveillance has been asked to do since the COVID-19 pandemic. The ‘nemmo’ R package makes this new approach conveniently usable by national and international surveillance teams, and we gratefully welcome comments and feedback on our proposal.

## Data Availability

All data produced are available online.

https://github.com/thlytras/analysis_nemmo_eurostat

https://github.com/thlytras/nemmo

---

**Supplementary Figure 1: Observed and expected mortality, Z-scores and P-scores between NeMMo and EuroMOMO, all countries**

**Supplementary Figure 2: Distribution of Z-scores and P-scores between NeMMo and EuroMOMO, all countries, entire time series**

## Notes

### Competing Interest Statement

The authors have declared no competing interest.

