## Supplementary Figure 1 for "NeMMo: an improved statistical algorithm for excess all-cause mortality surveillance and monitoring"

### Austria, observed and expected mortality

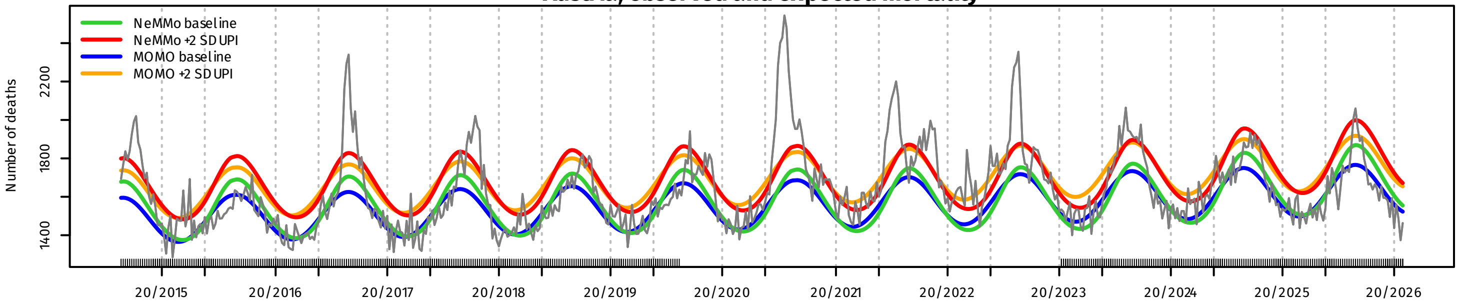


### Austria, Z-scores

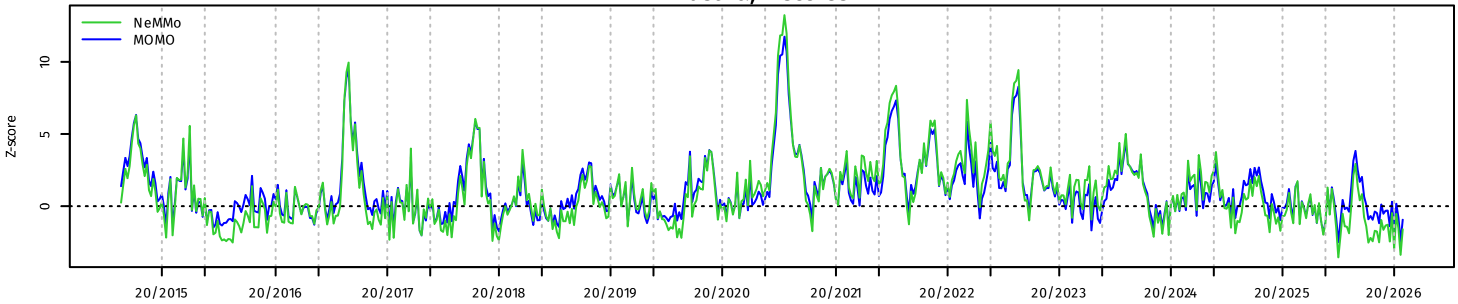


### Austria, P-scores

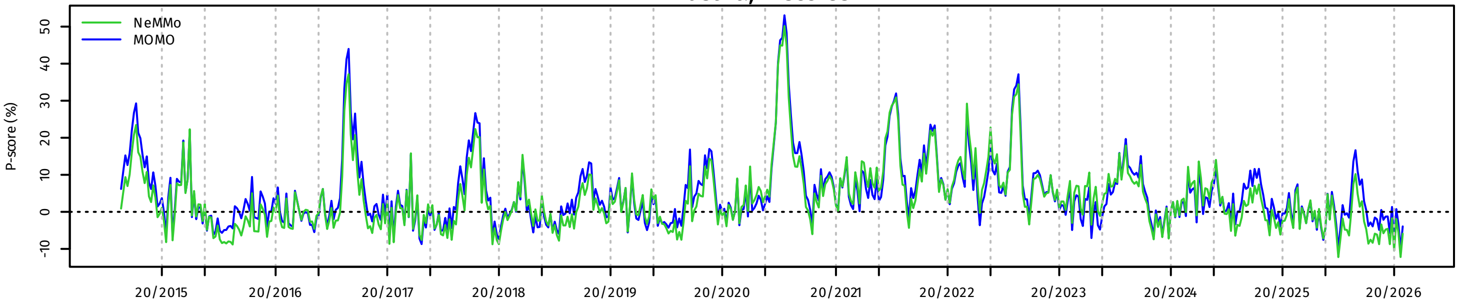


### Belgium, observed and expected mortality

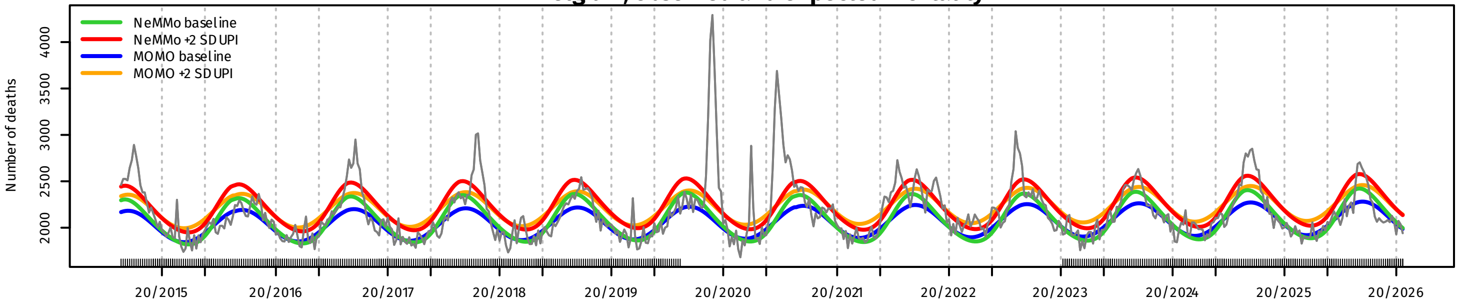


### Belgium, Z-scores

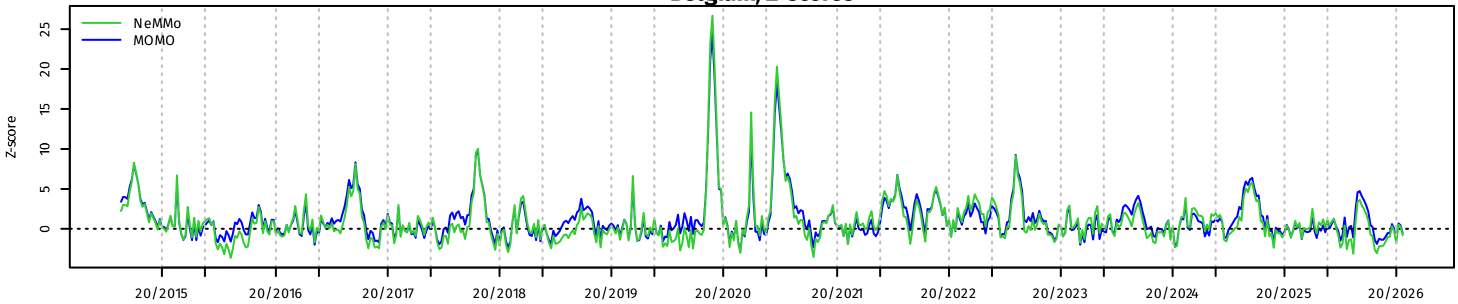


### Belgium, P-scores

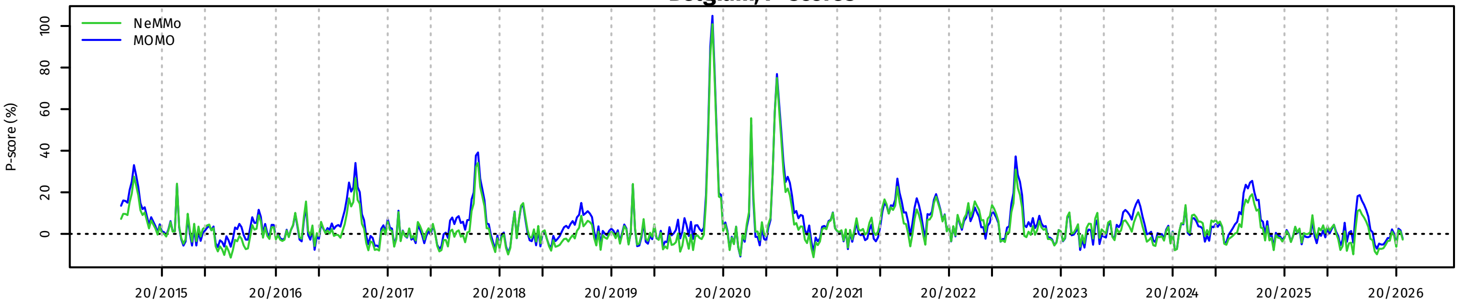

Bulgaria, observed and expected mortality

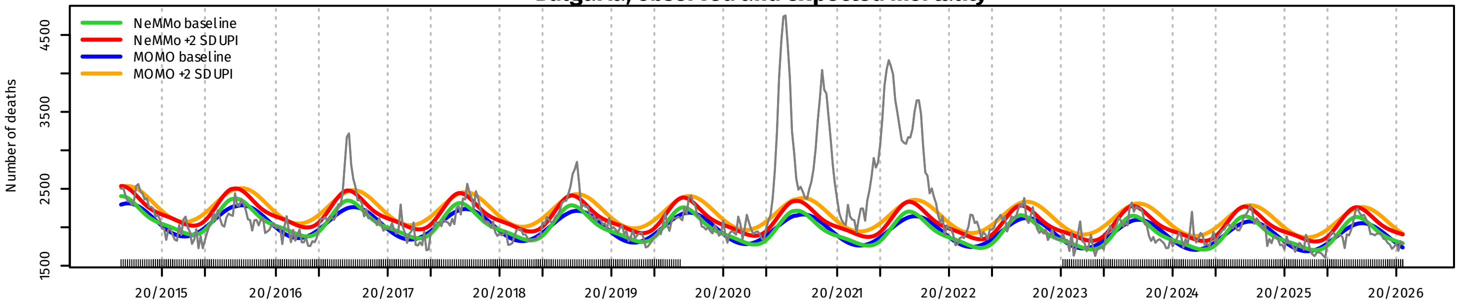


Bulgaria, Z-scores

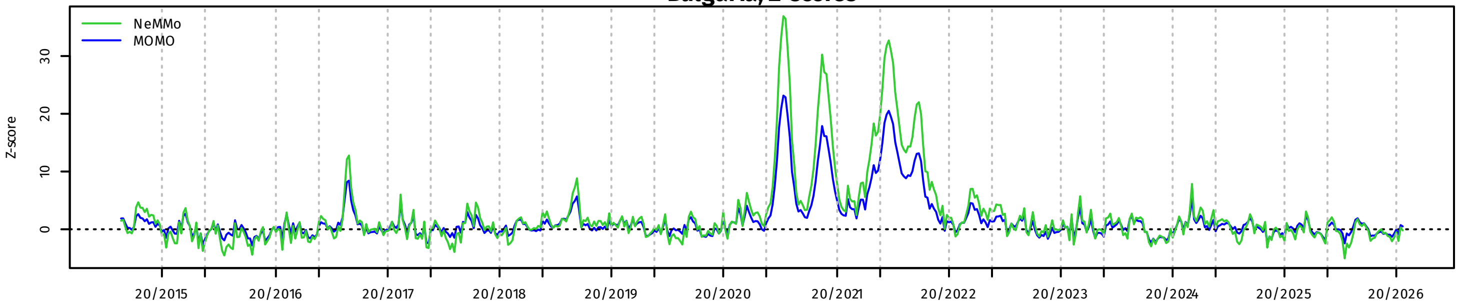


Bulgaria, P-scores

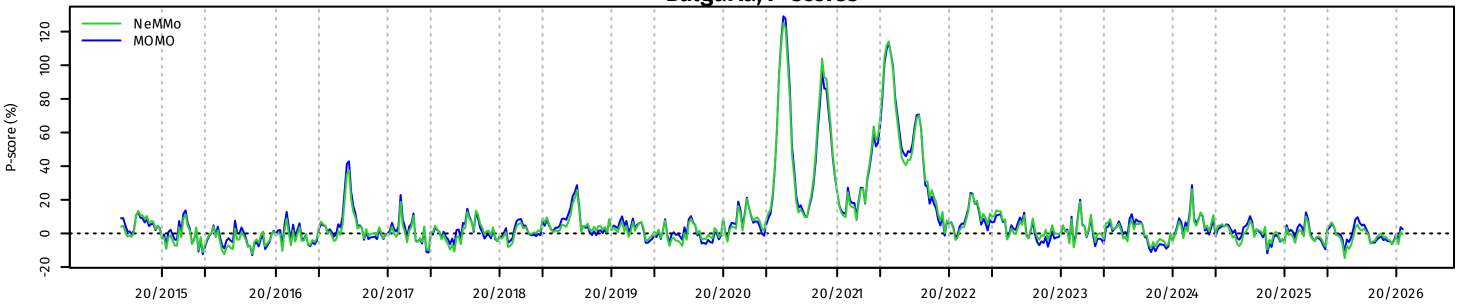


Croatia, observed and expected mortality

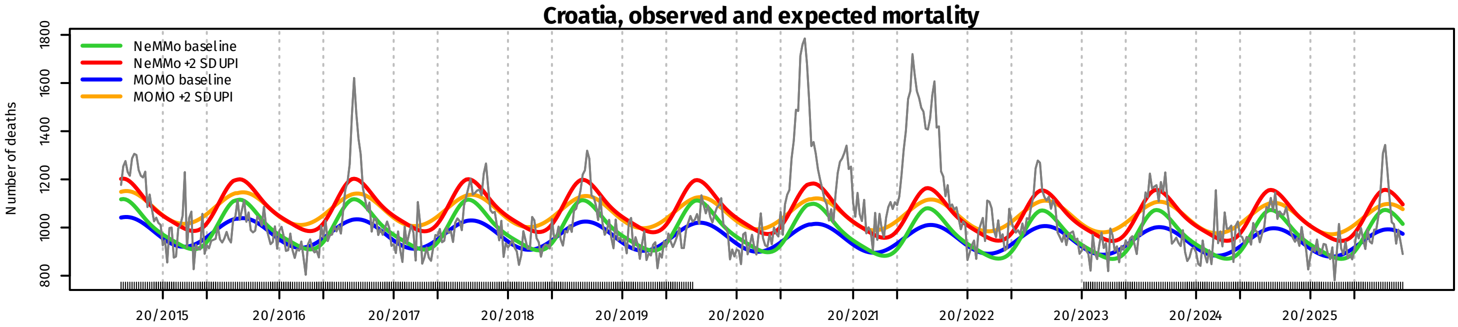


Croatia, Z-scores

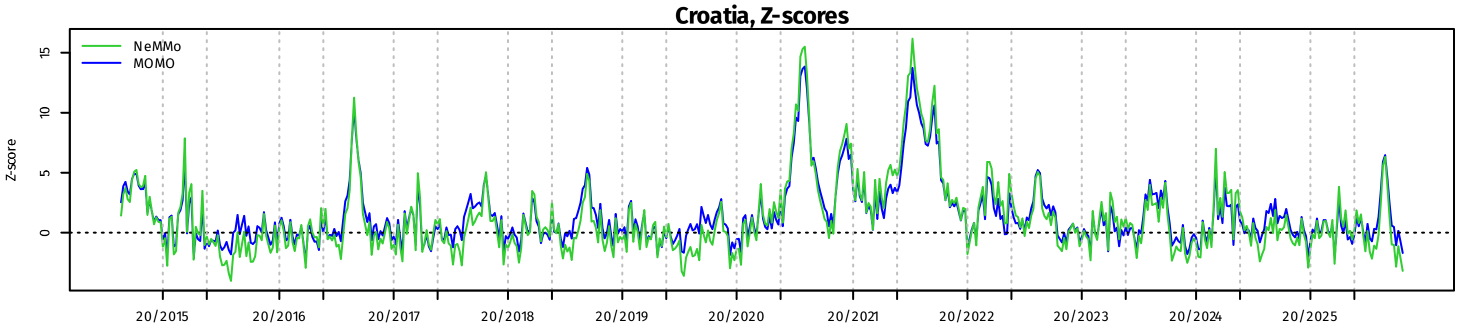


Croatia, P-scores

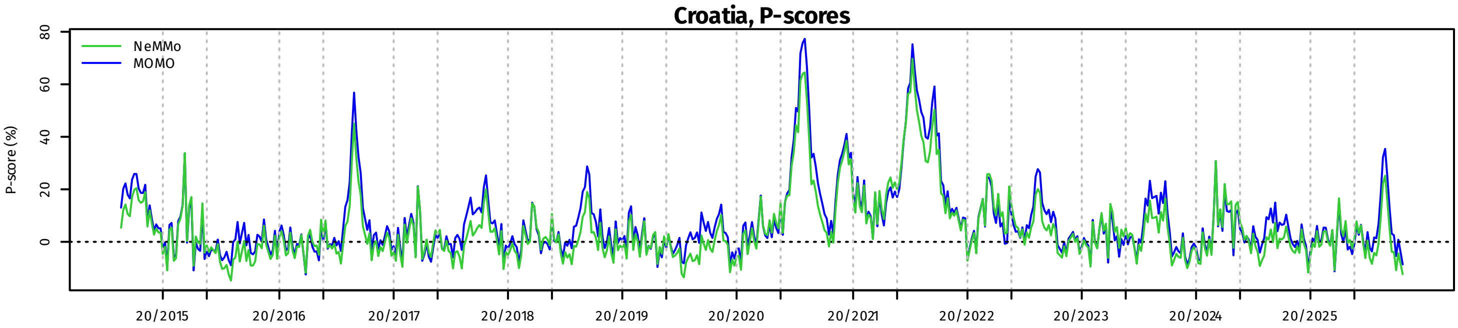

Cyprus, observed and expected mortality

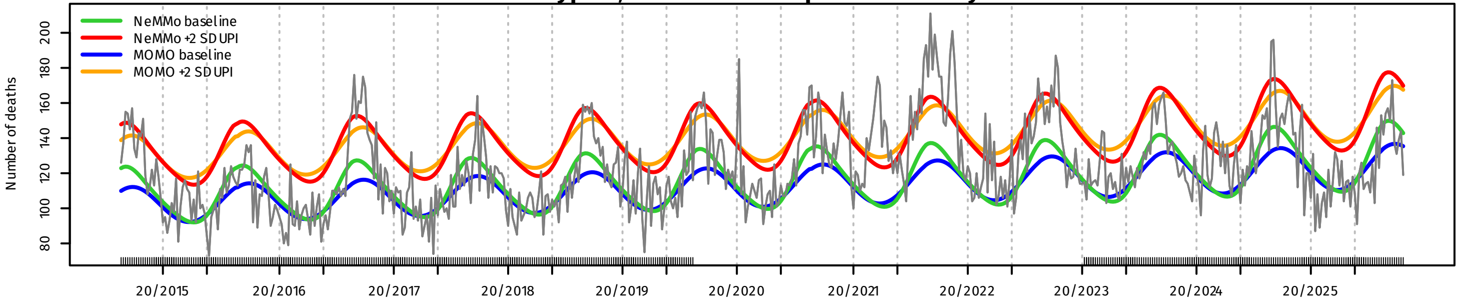


Cyprus, Z-scores

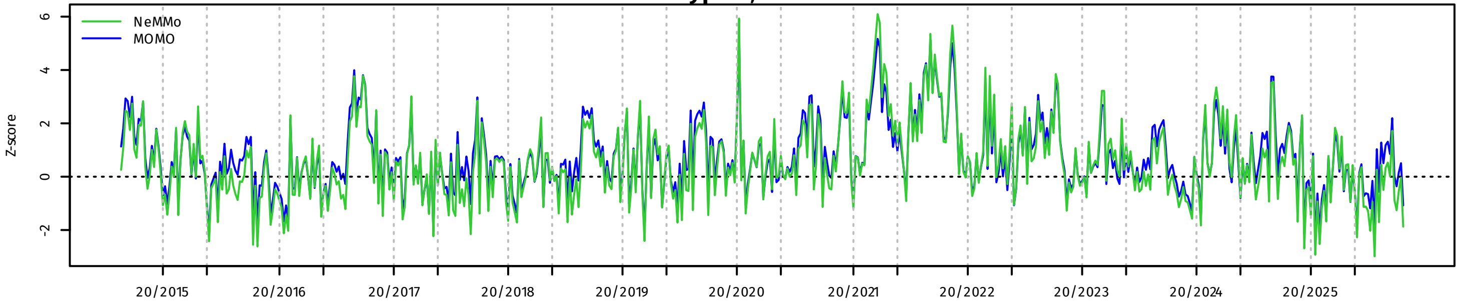


Cyprus, P-scores

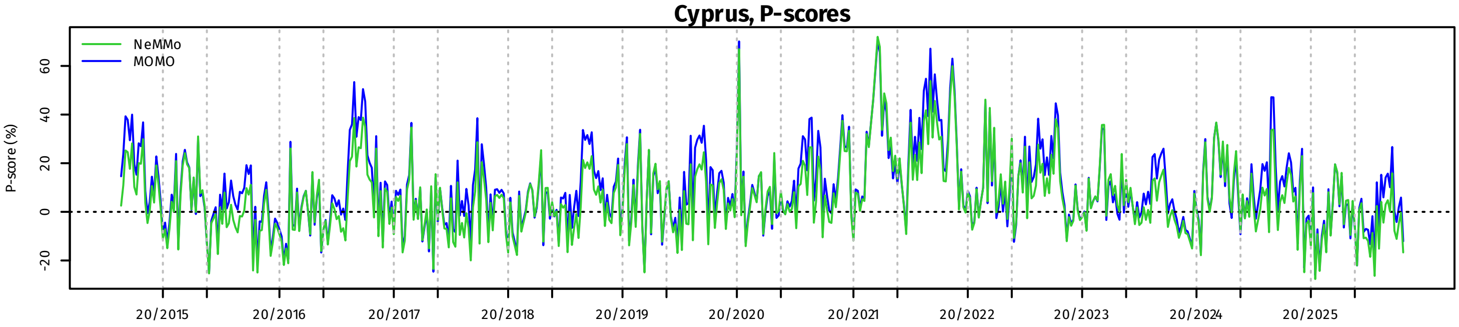


Czechia, observed and expected mortality

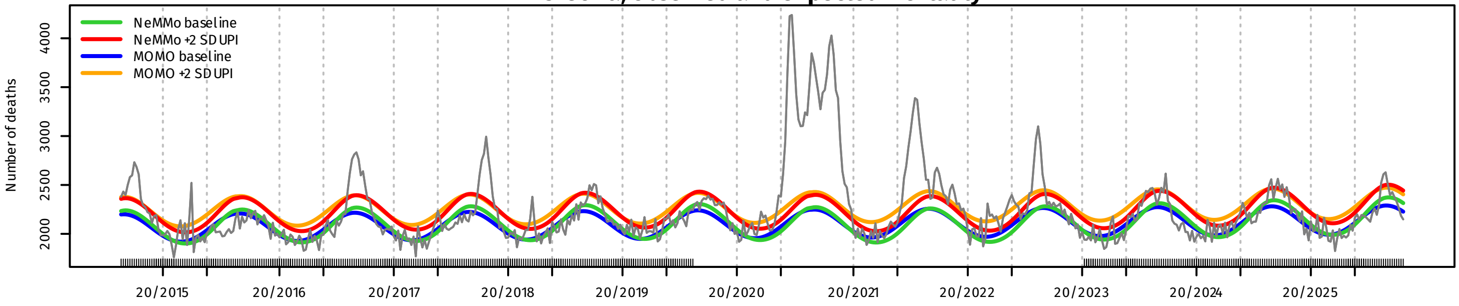


Czechia, Z-scores

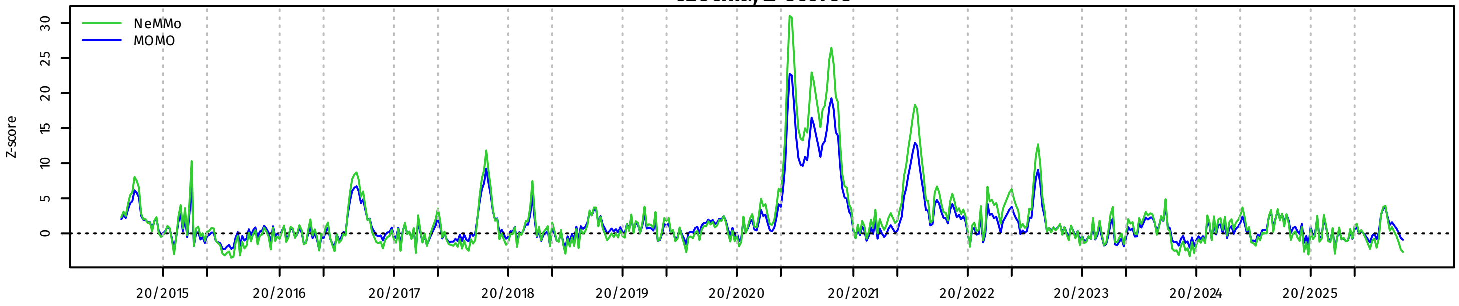


Czechia, P-scores

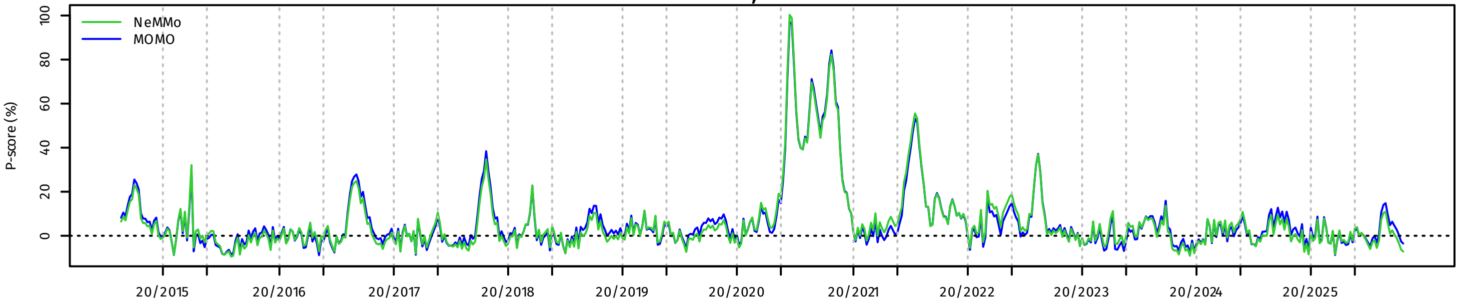

**Denmark, observed and expected mortality**

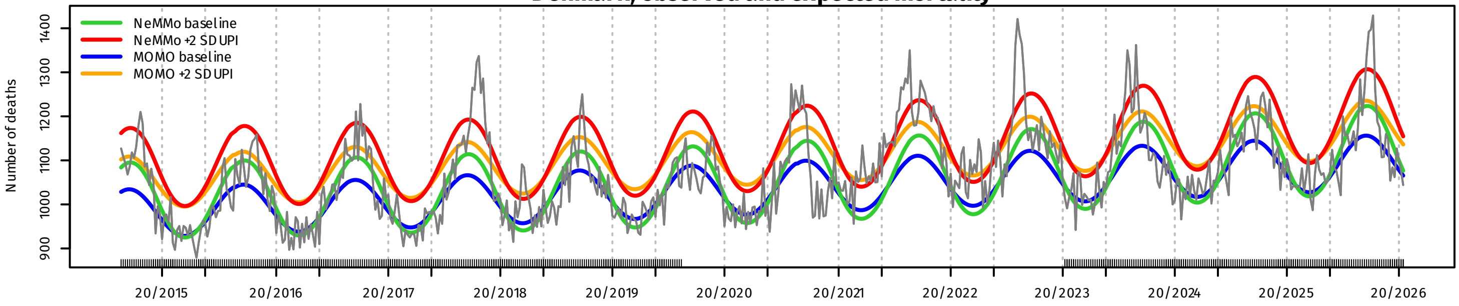


**Denmark, Z-scores**

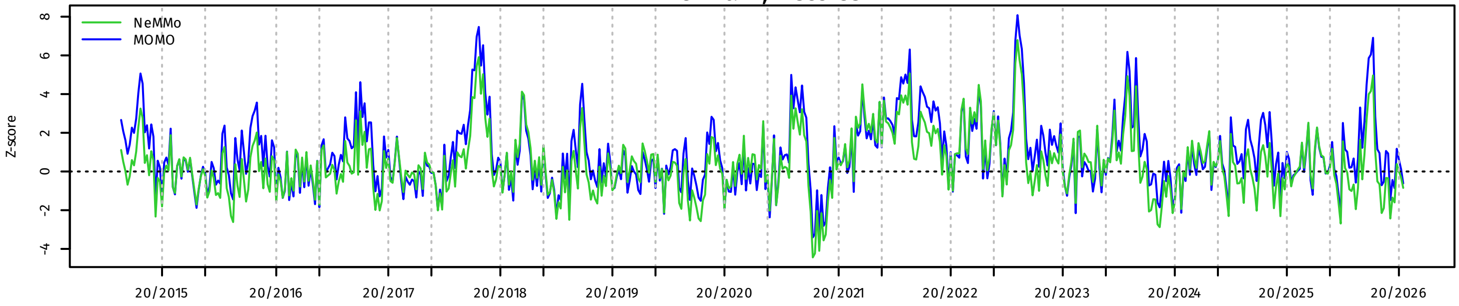


**Denmark, P-scores**

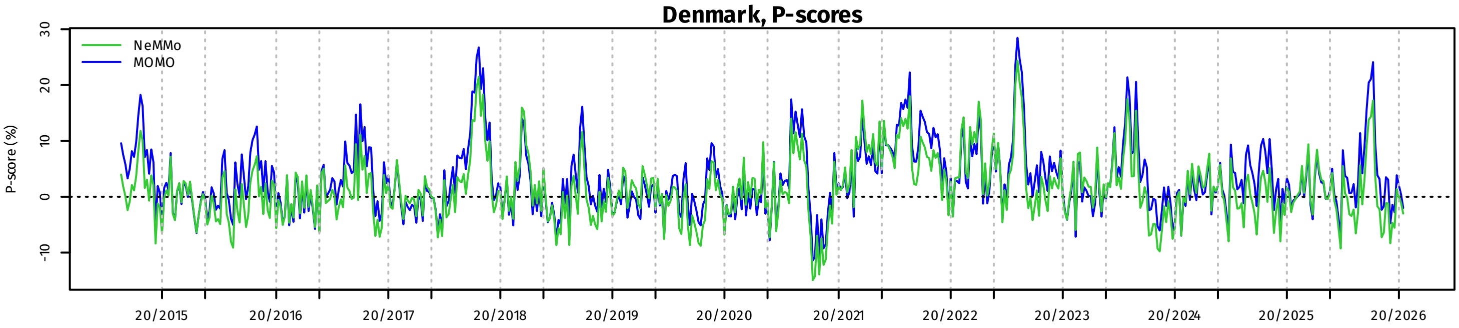


**Estonia, observed and expected mortality**

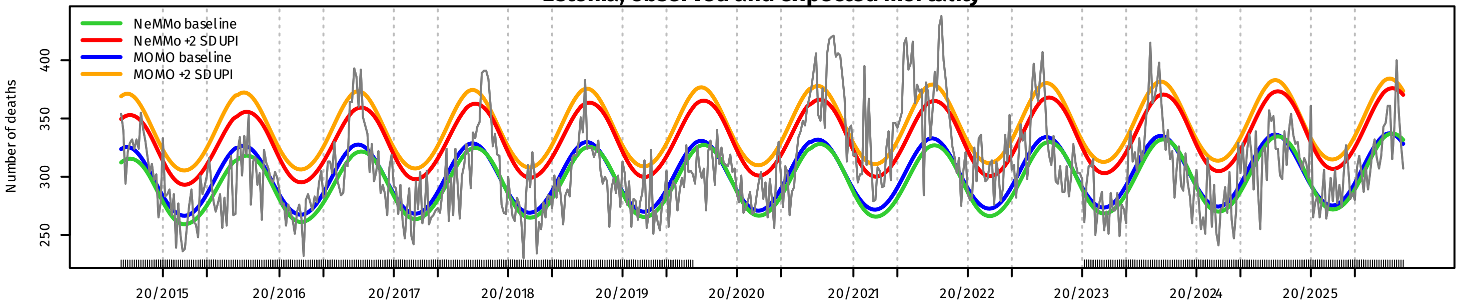


**Estonia, Z-scores**

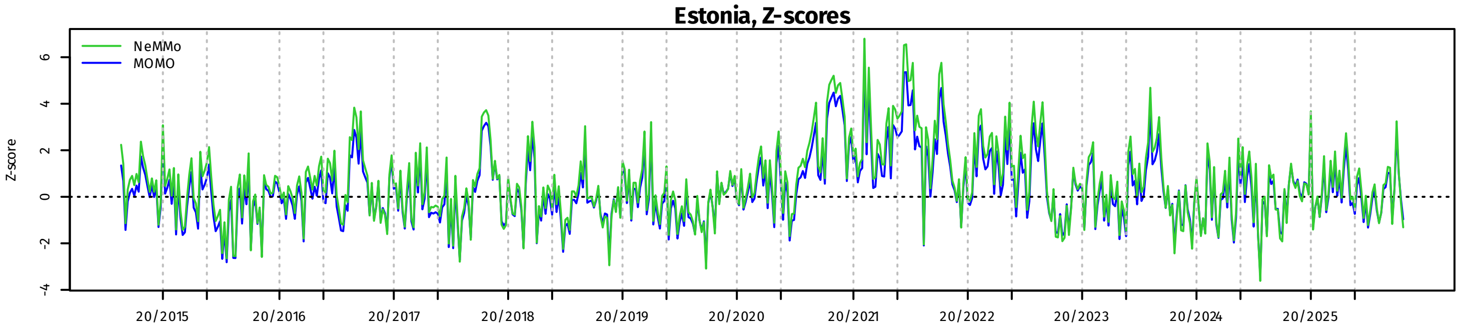


**Estonia, P-scores**

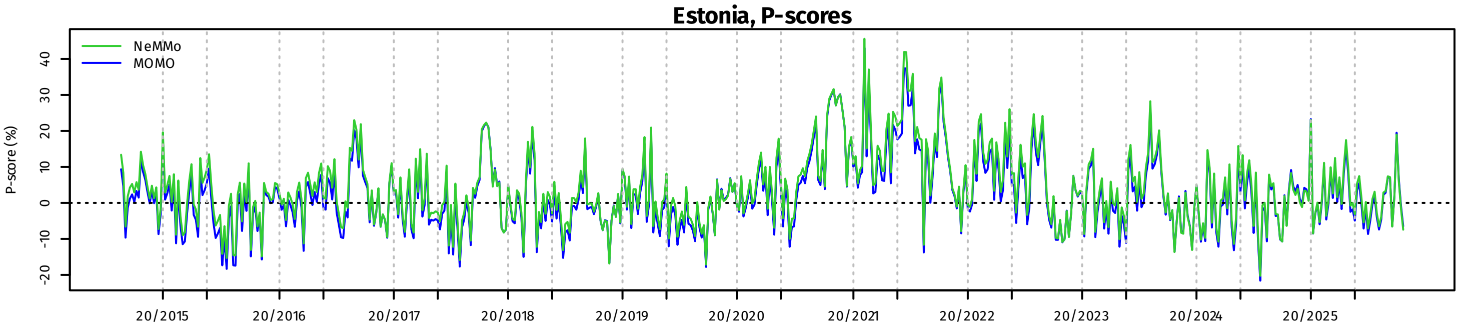

**Finland, observed and expected mortality**

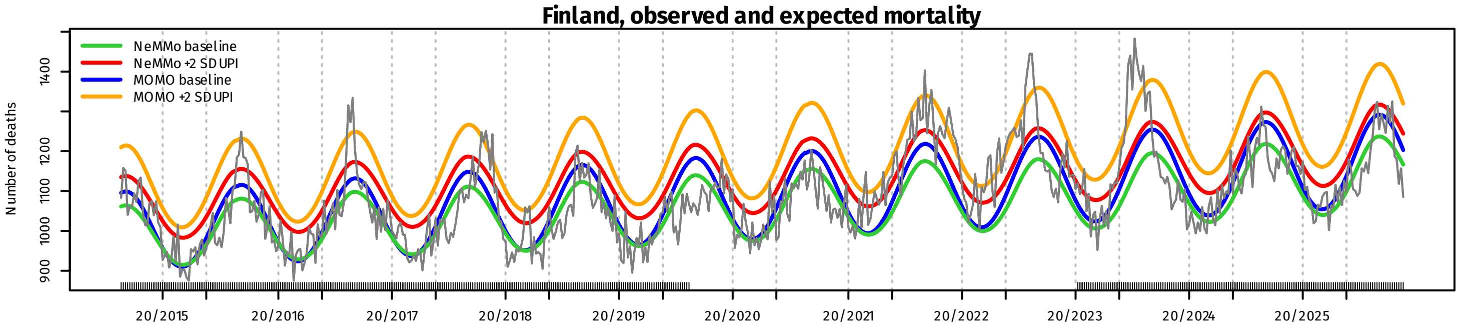


**Finland, Z-scores**

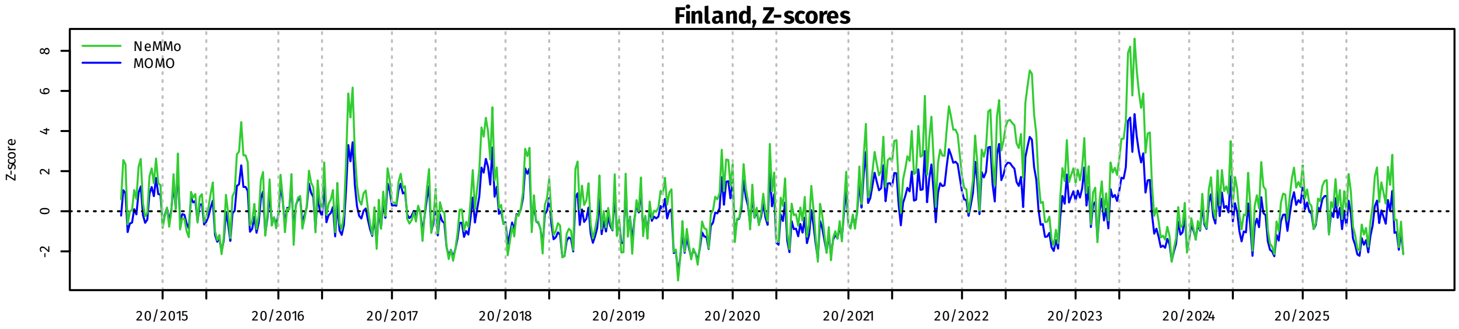


**Finland, P-scores**

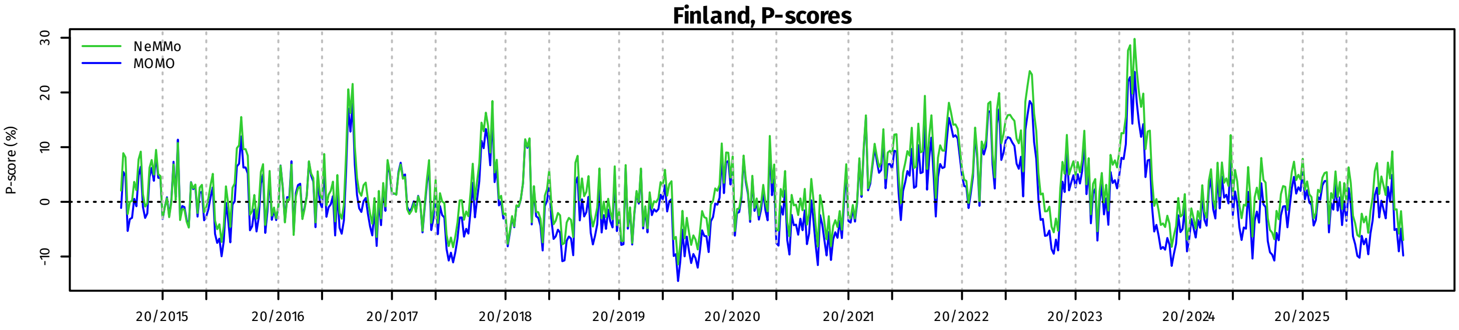


**France, observed and expected mortality**

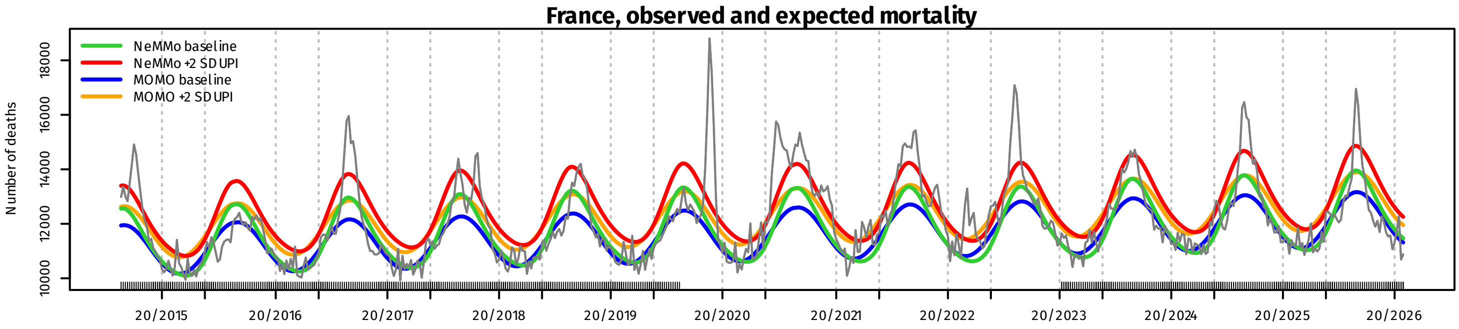


**France, Z-scores**

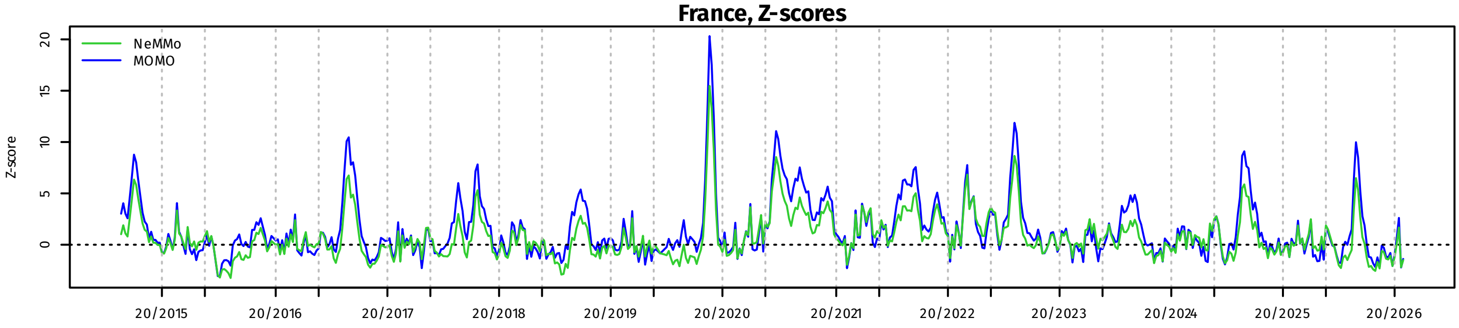


**France, P-scores**

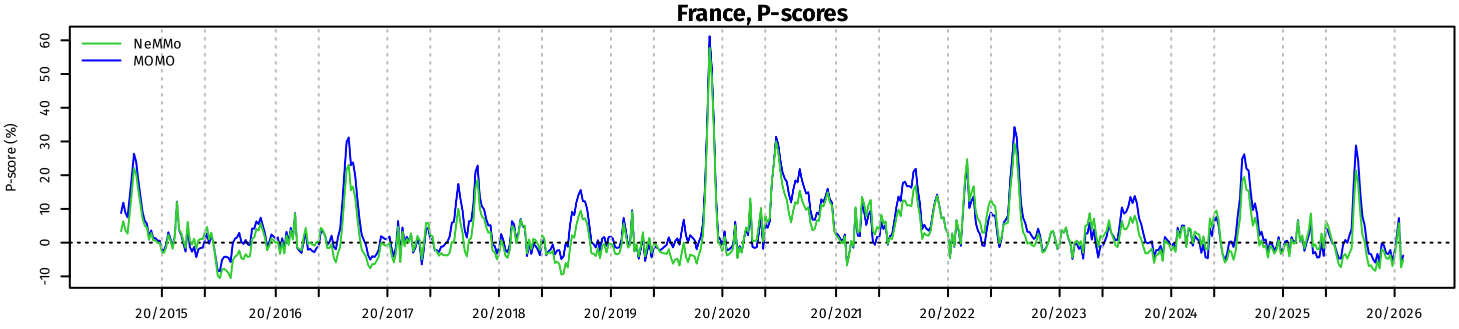

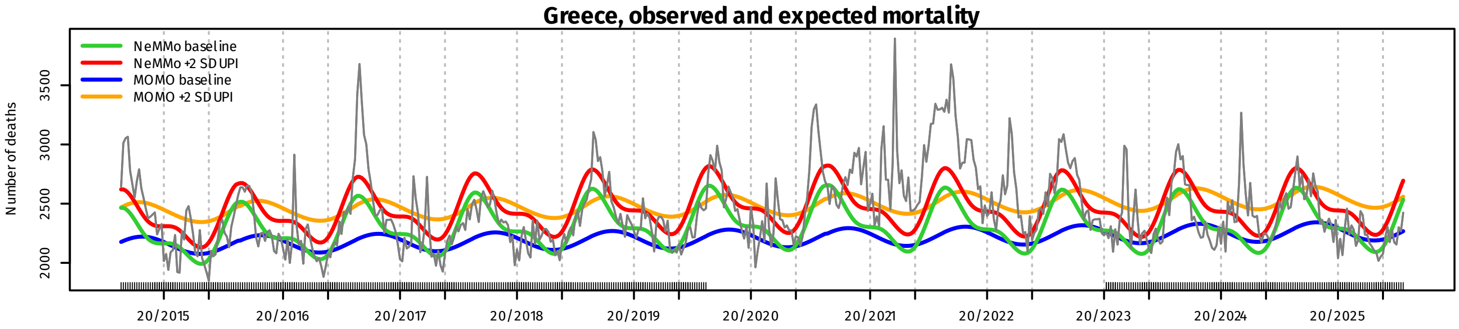
Greece, observed and expected mortality
NeMMo baseline
NeMMo +2 SD UPI
MOMO baseline
MOMO +2 SD UPI
Number of deaths
2000
2500
3000
3500
20/2015
20/2016
20/2017
20/2018
20/2019
20/2020
20/2021
20/2022
20/2023
20/2024
20/2025

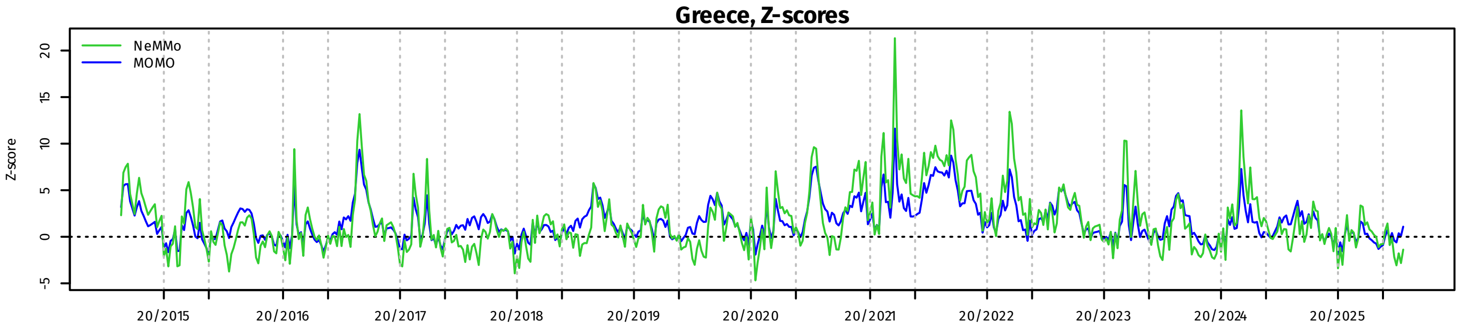
Greece, Z-scores
NeMMo
MOMO
Z-score
-5
0
5
10
15
20
20/2015
20/2016
20/2017
20/2018
20/2019
20/2020
20/2021
20/2022
20/2023
20/2024
20/2025

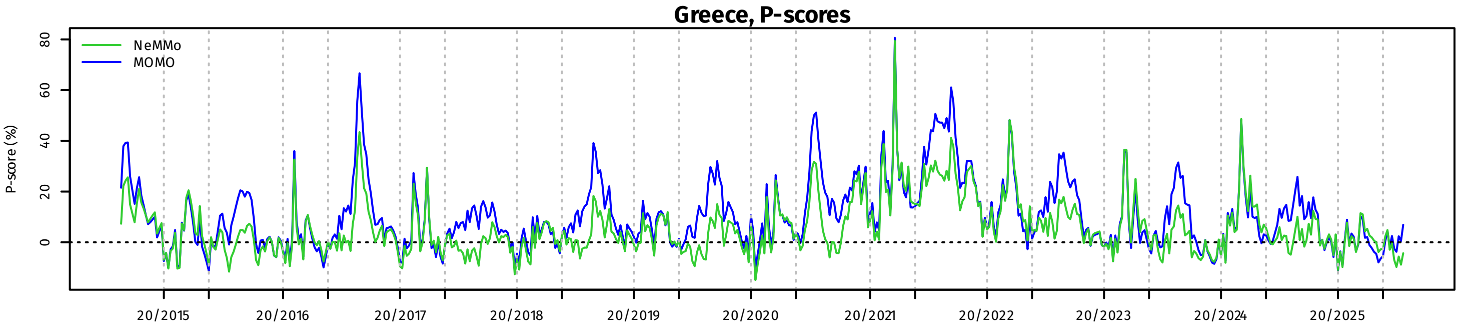
Greece, P-scores
NeMMo
MOMO
P-score (%)
0
20
40
60
80
20/2015
20/2016
20/2017
20/2018
20/2019
20/2020
20/2021
20/2022
20/2023
20/2024
20/2025

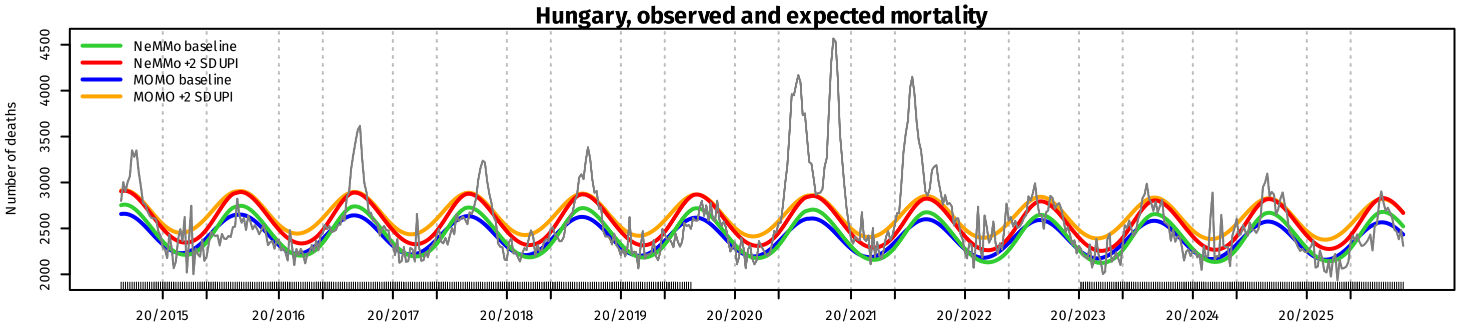
Hungary, observed and expected mortality
NeMMo baseline
NeMMo +2 SD UPI
MOMO baseline
MOMO +2 SD UPI
Number of deaths
2000
2500
3000
3500
4000
4500
20/2015
20/2016
20/2017
20/2018
20/2019
20/2020
20/2021
20/2022
20/2023
20/2024
20/2025

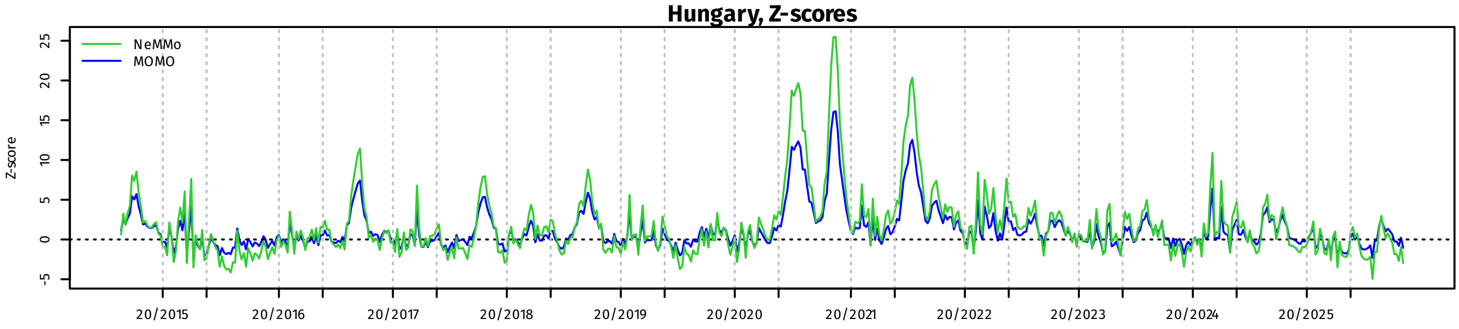
Hungary, Z-scores
NeMMo
MOMO
Z-score
-5
0
5
10
15
20
25
20/2015
20/2016
20/2017
20/2018
20/2019
20/2020
20/2021
20/2022
20/2023
20/2024
20/2025

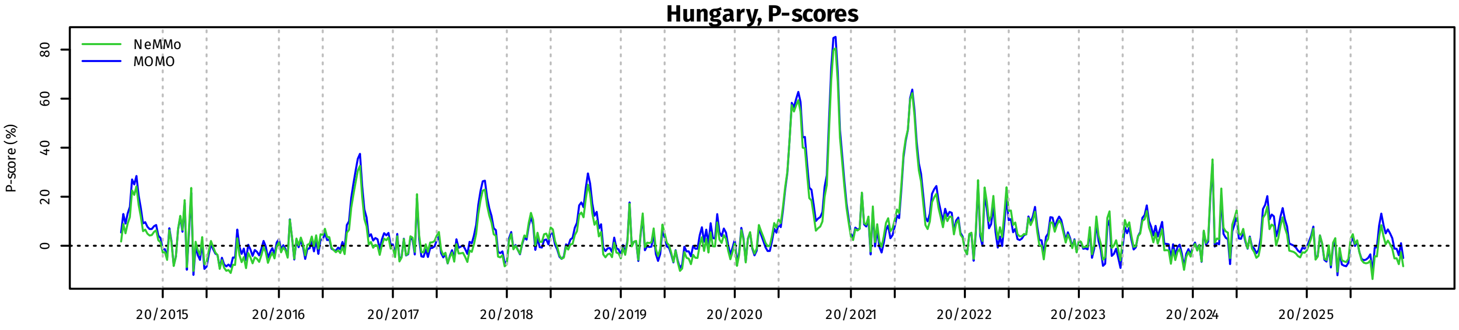
Hungary, P-scores
NeMMo
MOMO
P-score (%)
0
20
40
60
80
20/2015
20/2016
20/2017
20/2018
20/2019
20/2020
20/2021
20/2022
20/2023
20/2024
20/2025

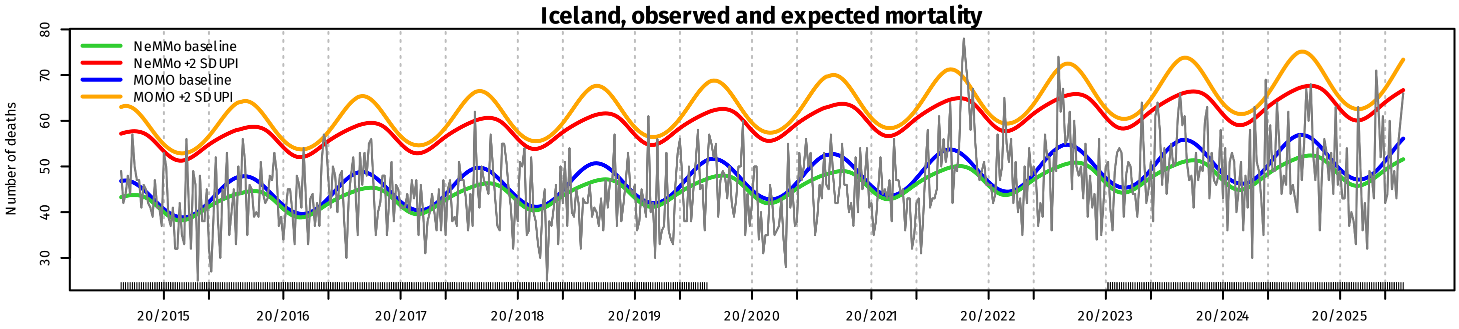
Iceland, observed and expected mortality
NeMMo baseline
NeMMo +2 SD UPI
MOMO baseline
MOMO +2 SD UPI
Number of deaths
30
40
50
60
70
80
20/2015
20/2016
20/2017
20/2018
20/2019
20/2020
20/2021
20/2022
20/2023
20/2024
20/2025

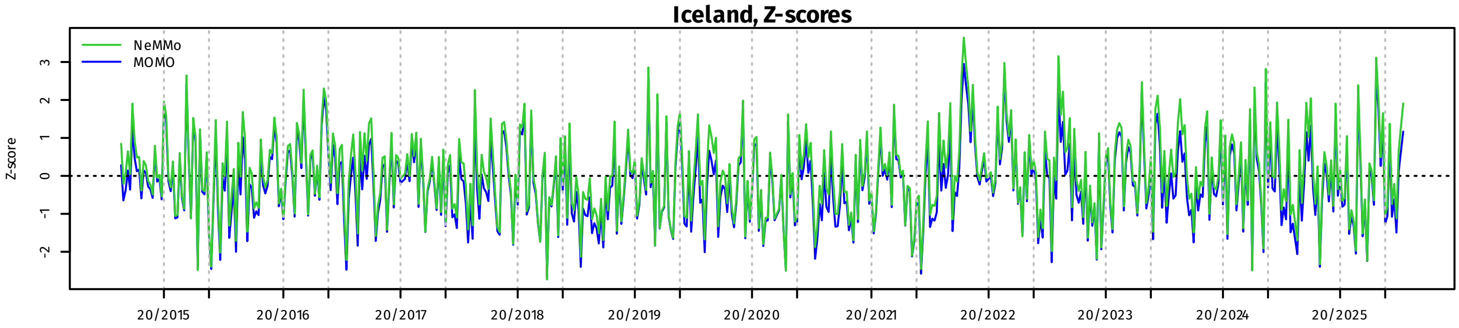
Iceland, Z-scores
NeMMo
MOMO
Z-score
-2
-1
0
1
2
3
20/2015
20/2016
20/2017
20/2018
20/2019
20/2020
20/2021
20/2022
20/2023
20/2024
20/2025

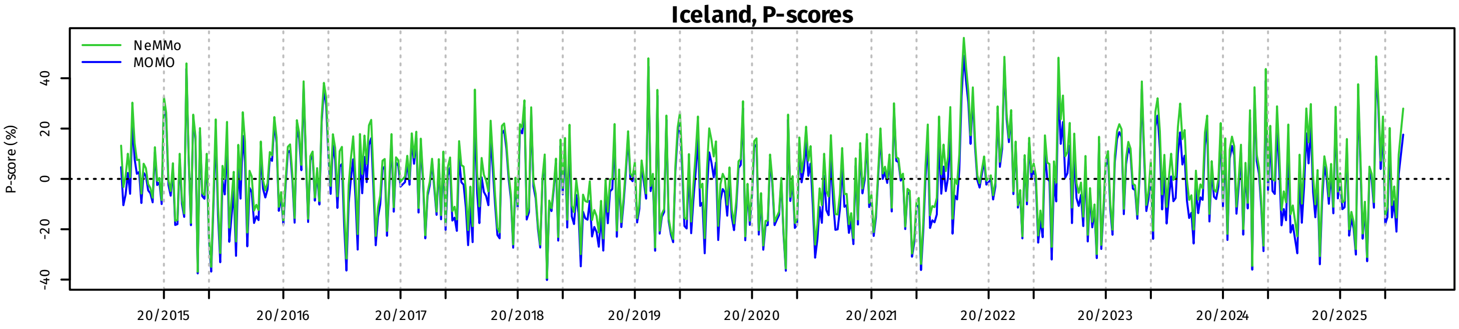
Iceland, P-scores
NeMMo
MOMO
P-score (%)
-40
-20
0
20
40
20/2015
20/2016
20/2017
20/2018
20/2019
20/2020
20/2021
20/2022
20/2023
20/2024
20/2025

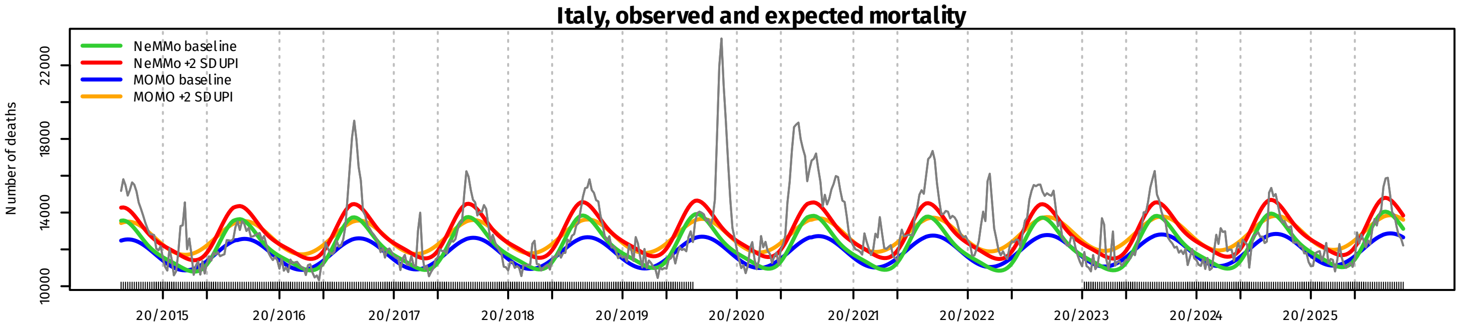
Italy, observed and expected mortality
NeMMo baseline
NeMMo +2 SD UPI
MOMO baseline
MOMO +2 SD UPI
Number of deaths
10000
14000
18000
22000
20/2015
20/2016
20/2017
20/2018
20/2019
20/2020
20/2021
20/2022
20/2023
20/2024
20/2025

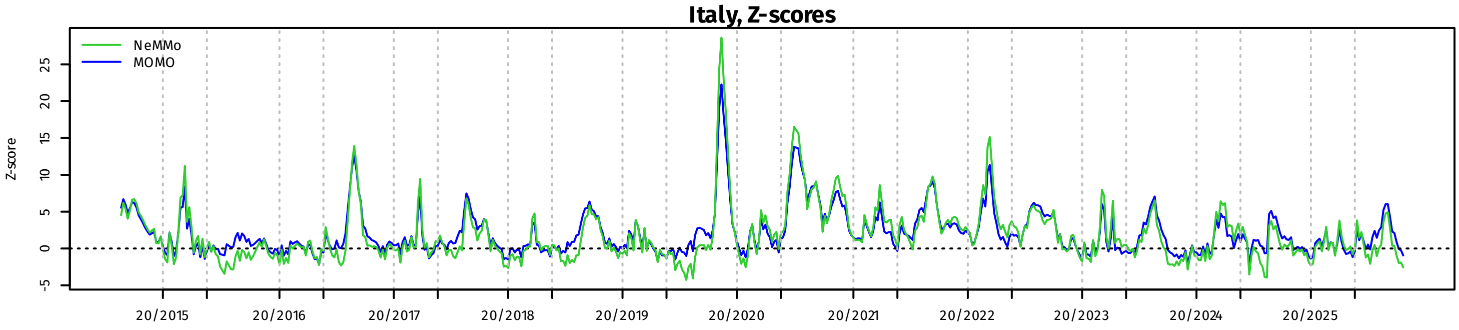
Italy, Z-scores
NeMMo
MOMO
Z-score
-5
0
5
10
15
20
25
20/2015
20/2016
20/2017
20/2018
20/2019
20/2020
20/2021
20/2022
20/2023
20/2024
20/2025

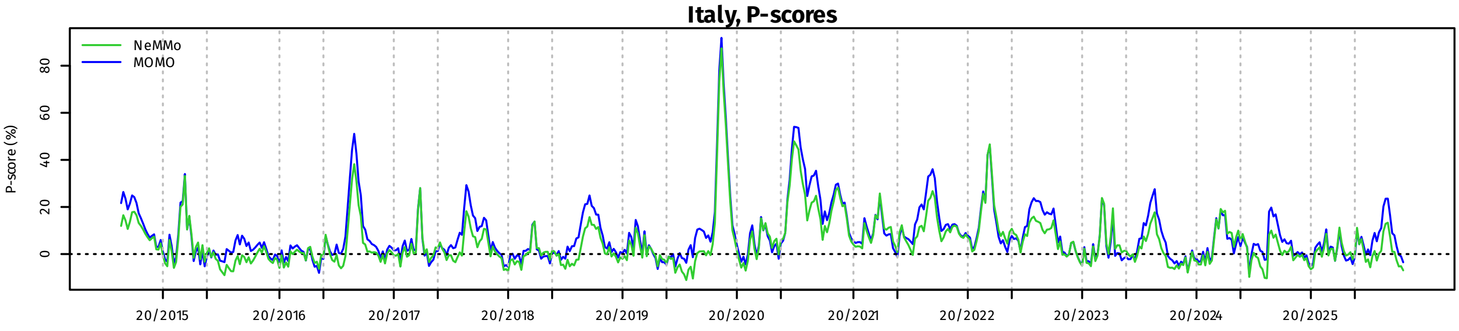
Italy, P-scores
NeMMo
MOMO
P-score (%)
0
20
40
60
80
20/2015
20/2016
20/2017
20/2018
20/2019
20/2020
20/2021
20/2022
20/2023
20/2024
20/2025

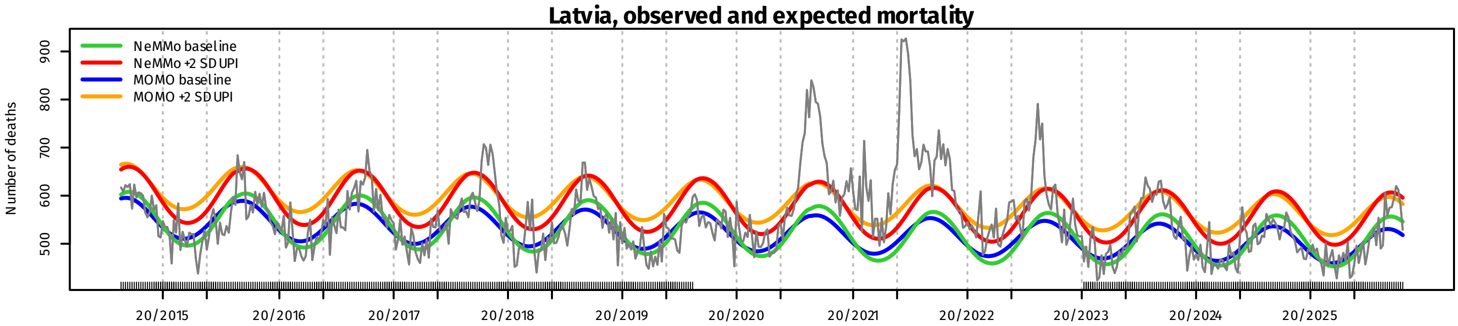
Latvia, observed and expected mortality
NeMMo baseline
NeMMo +2 SD UPI
MOMO baseline
MOMO +2 SD UPI
Number of deaths
500
600
700
800
900
20/2015
20/2016
20/2017
20/2018
20/2019
20/2020
20/2021
20/2022
20/2023
20/2024
20/2025

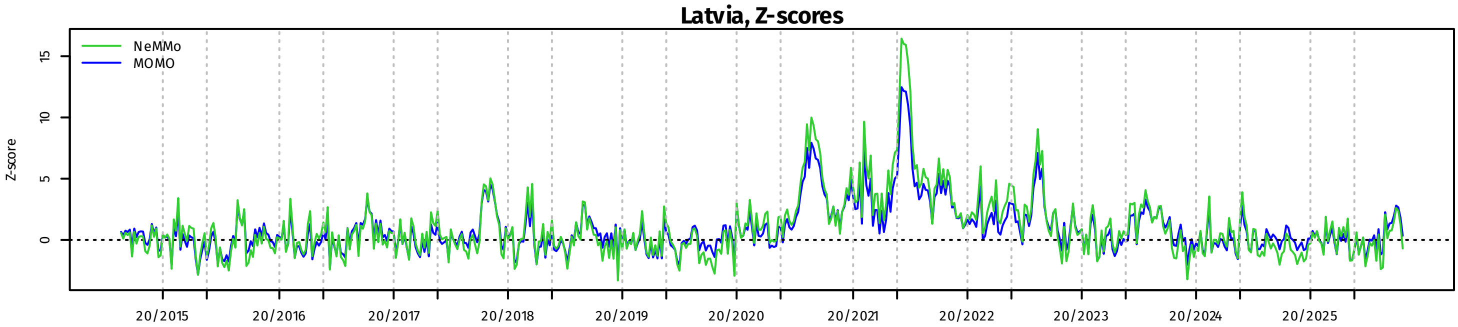
Latvia, Z-scores
NeMMo
MOMO
Z-score
0
5
10
15
20/2015
20/2016
20/2017
20/2018
20/2019
20/2020
20/2021
20/2022
20/2023
20/2024
20/2025

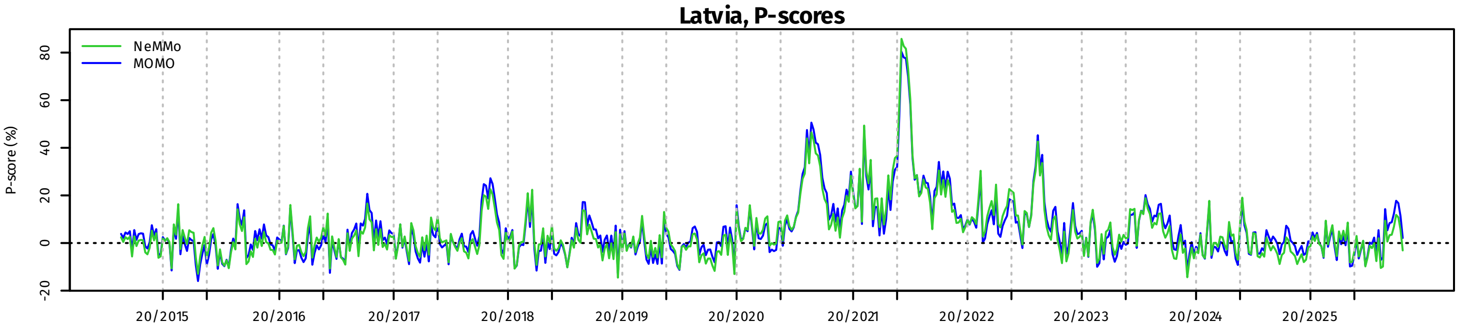
Latvia, P-scores
NeMMo
MOMO
P-score (%)
-20
0
20
40
60
80
20/2015
20/2016
20/2017
20/2018
20/2019
20/2020
20/2021
20/2022
20/2023
20/2024
20/2025

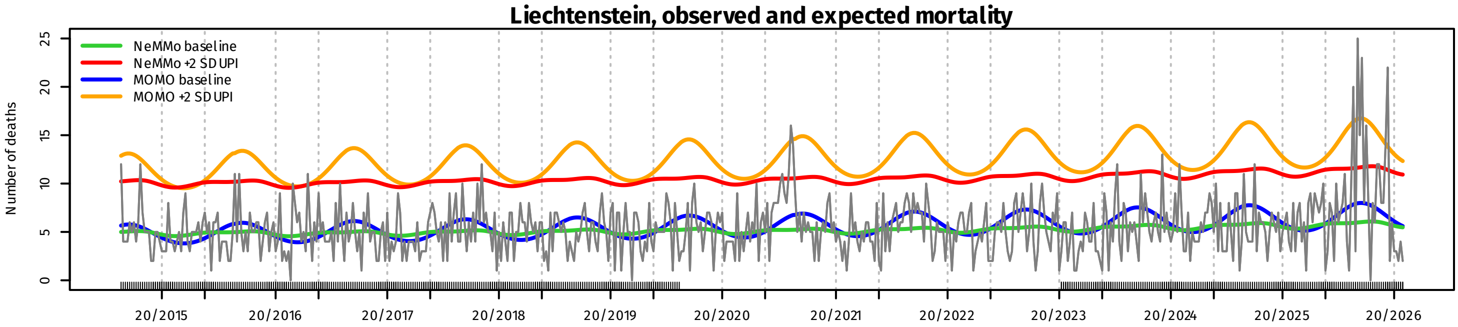
Liechtenstein, observed and expected mortality
NeMMo baseline
NeMMo +2 SD UPI
MOMO baseline
MOMO +2 SD UPI
Number of deaths
0
5
10
15
20
25
20/2015
20/2016
20/2017
20/2018
20/2019
20/2020
20/2021
20/2022
20/2023
20/2024
20/2025
20/2026

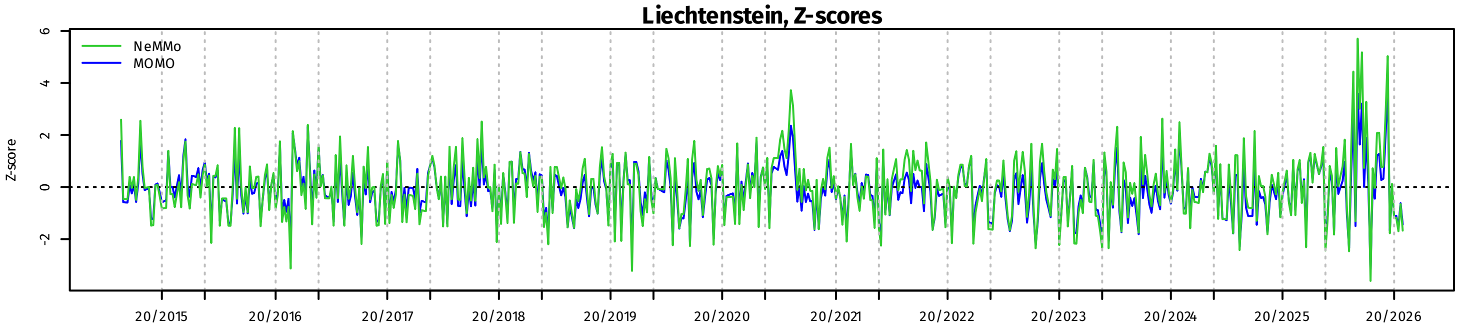
Liechtenstein, Z-scores
NeMMo
MOMO
Z-score
-2
0
2
4
6
20/2015
20/2016
20/2017
20/2018
20/2019
20/2020
20/2021
20/2022
20/2023
20/2024
20/2025
20/2026

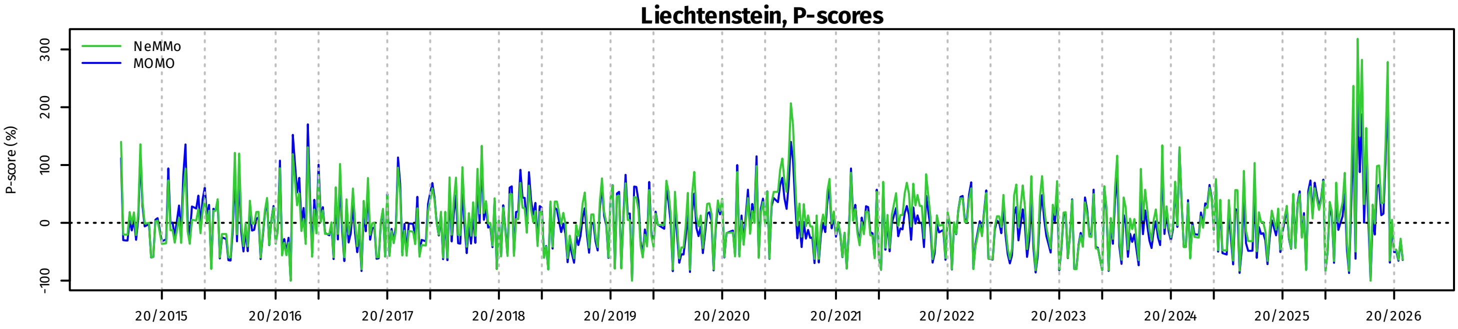
Liechtenstein, P-scores
NeMMo
MOMO
P-score (%)
-100
0
100
200
300
20/2015
20/2016
20/2017
20/2018
20/2019
20/2020
20/2021
20/2022
20/2023
20/2024
20/2025
20/2026

### Lithuania, observed and expected mortality

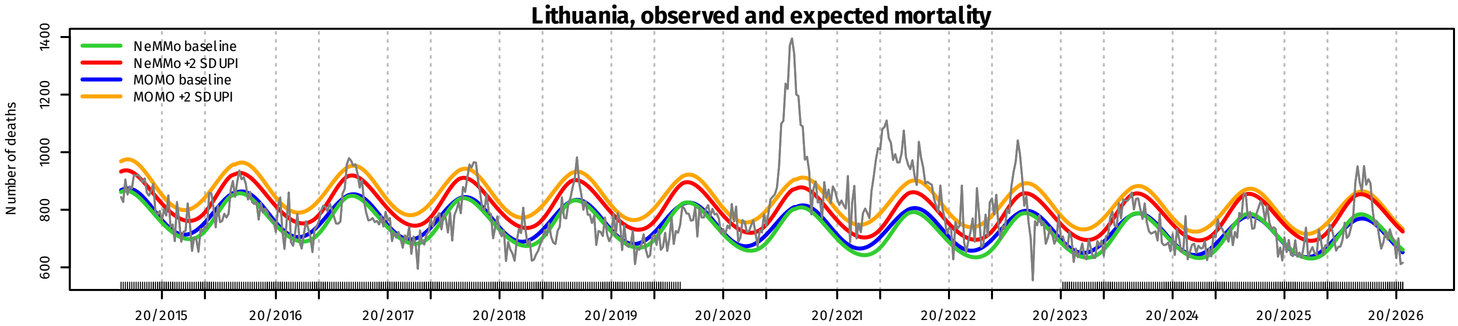


### Lithuania, Z-scores

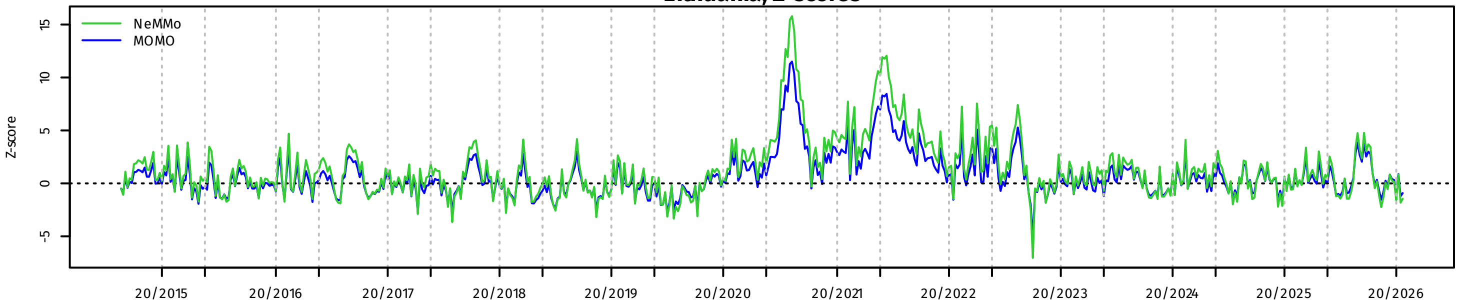


### Lithuania, P-scores

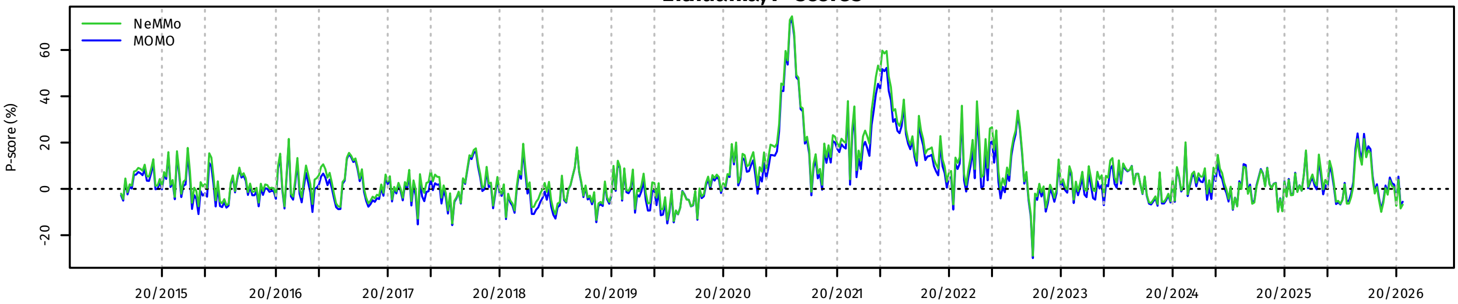


### Luxembourg, observed and expected mortality

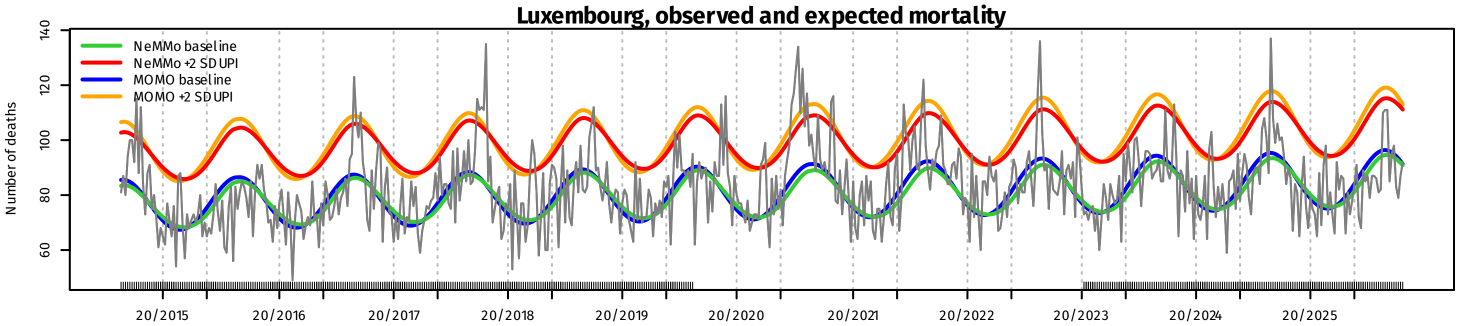


### Luxembourg, Z-scores

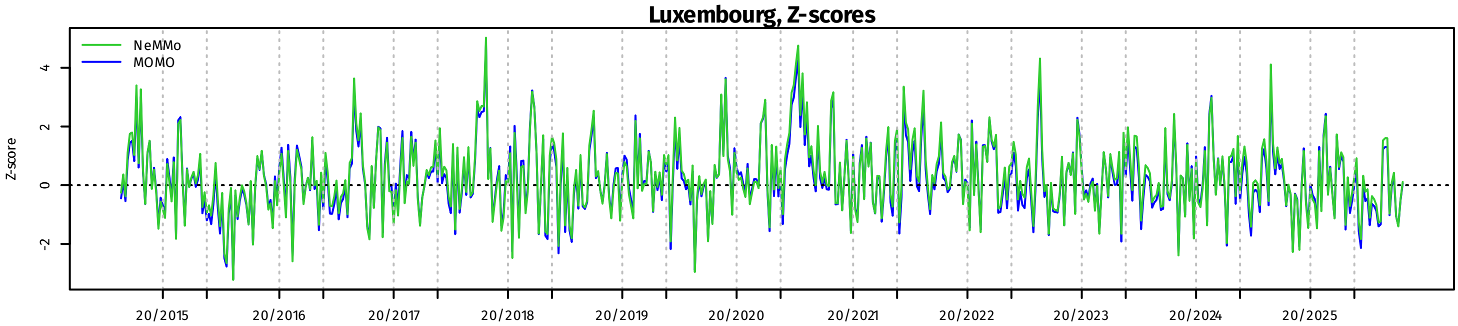


### Luxembourg, P-scores

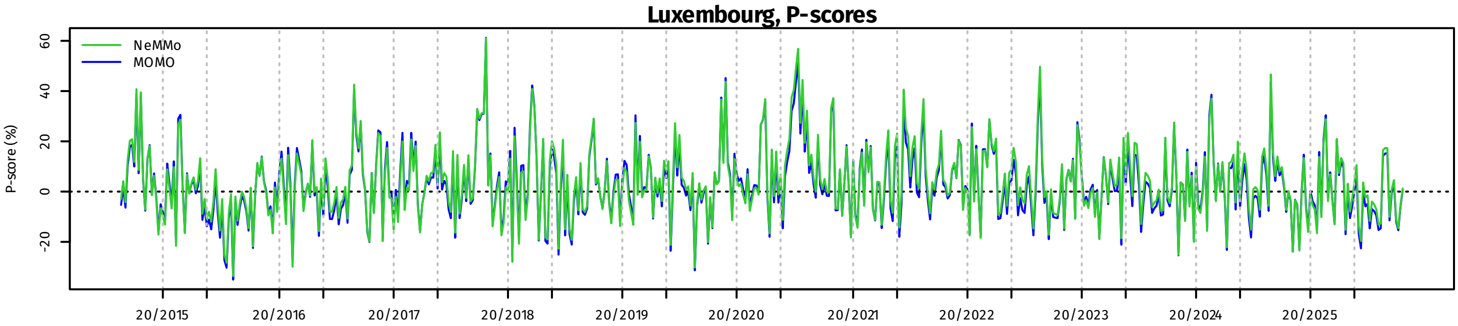

**Malta, observed and expected mortality**

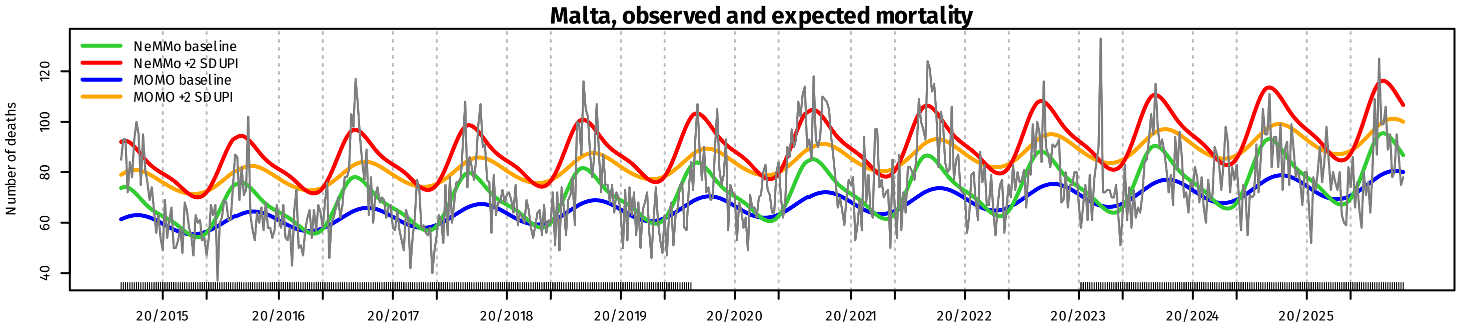


**Malta, Z-scores**

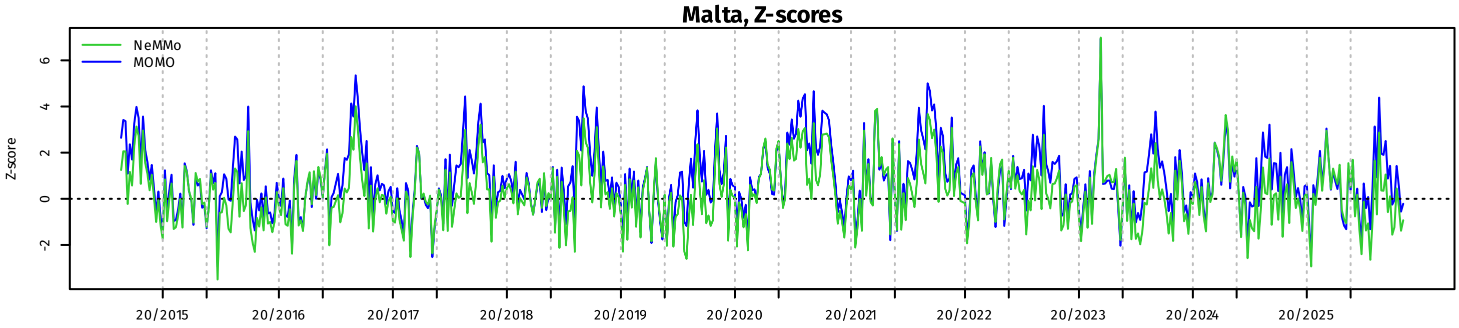


**Malta, P-scores**

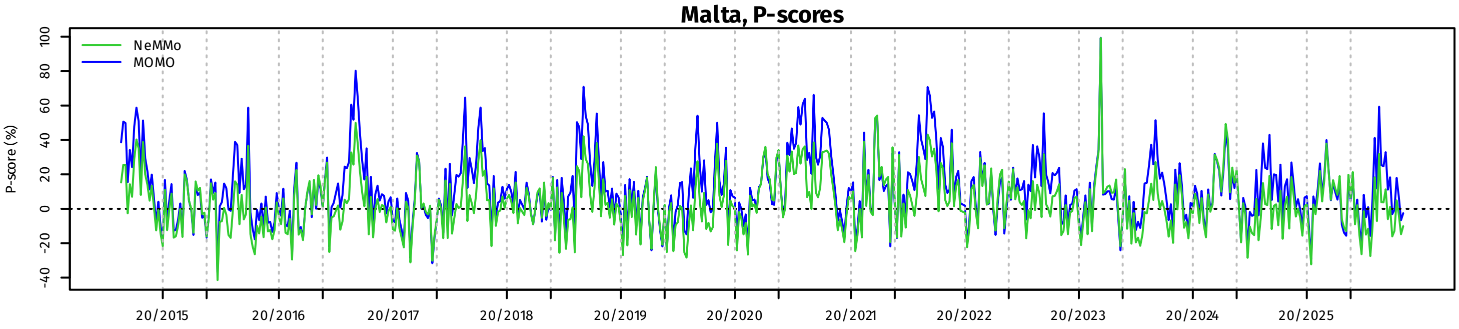


**Montenegro, observed and expected mortality**

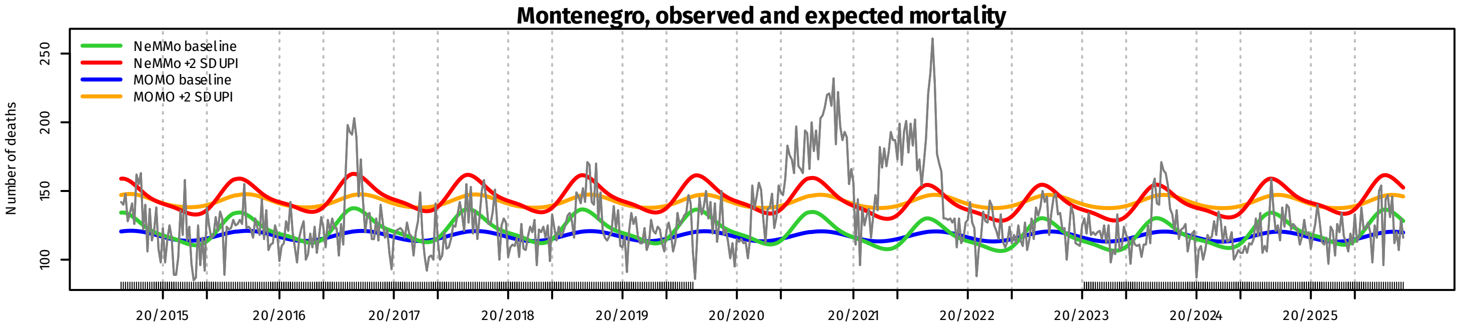


**Montenegro, Z-scores**

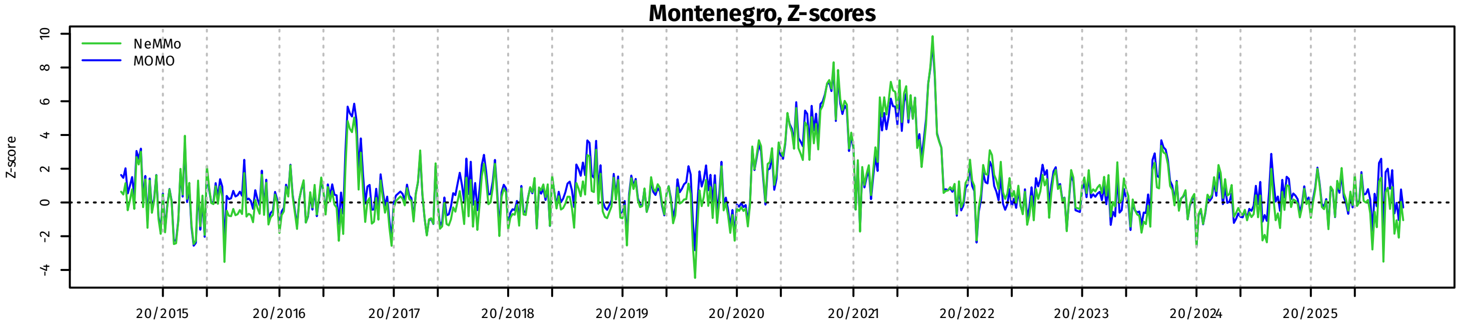


**Montenegro, P-scores**

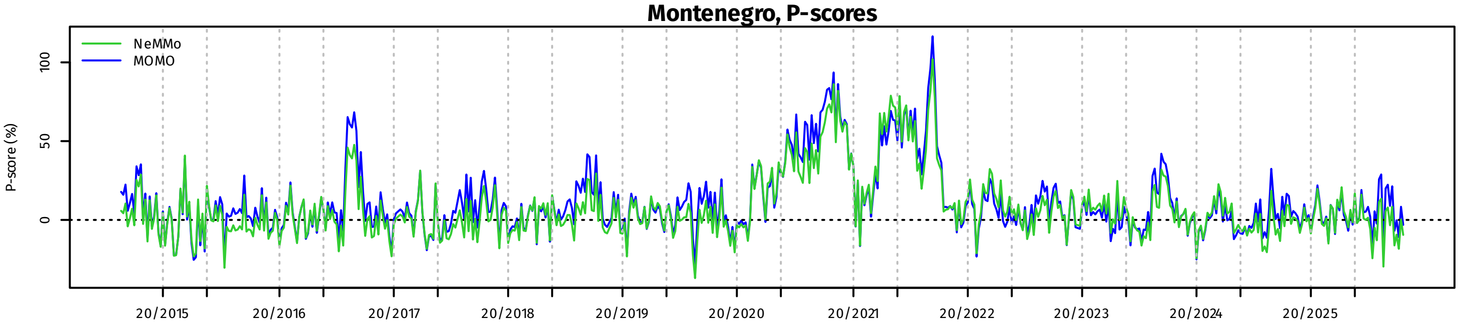

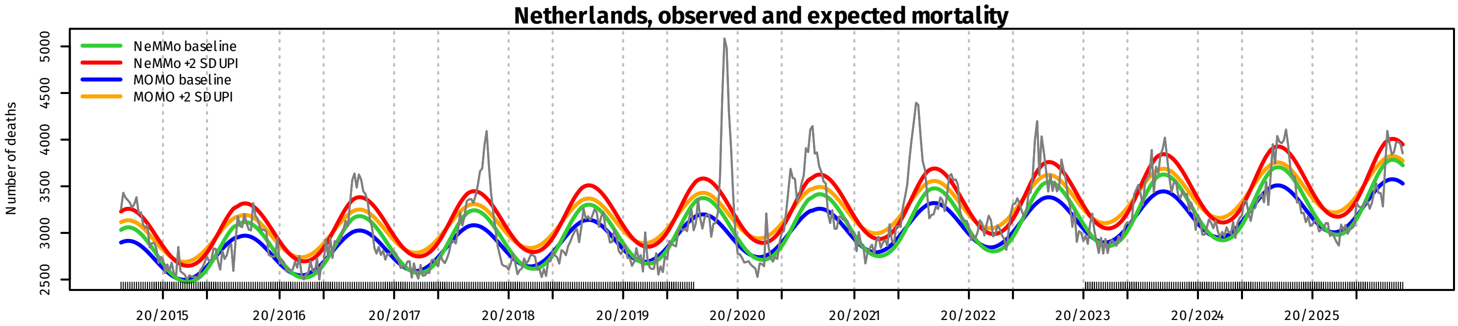
Netherlands, observed and expected mortality
NeMMo baseline
NeMMo +2 SD UPI
MOMO baseline
MOMO +2 SD UPI
Number of deaths
2500
3000
3500
4000
4500
5000
20/2015
20/2016
20/2017
20/2018
20/2019
20/2020
20/2021
20/2022
20/2023
20/2024
20/2025

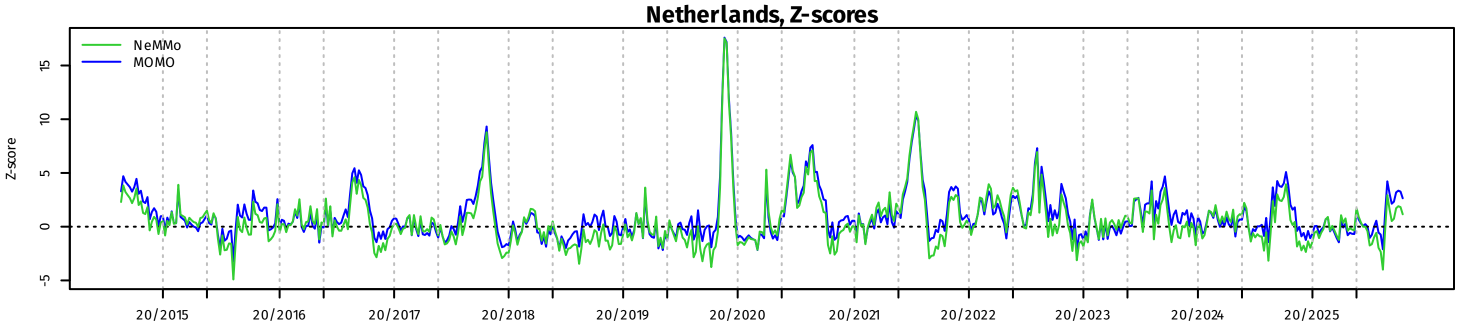
Netherlands, Z-scores
NeMMo
MOMO
Z-score
-5
0
5
10
15
20/2015
20/2016
20/2017
20/2018
20/2019
20/2020
20/2021
20/2022
20/2023
20/2024
20/2025

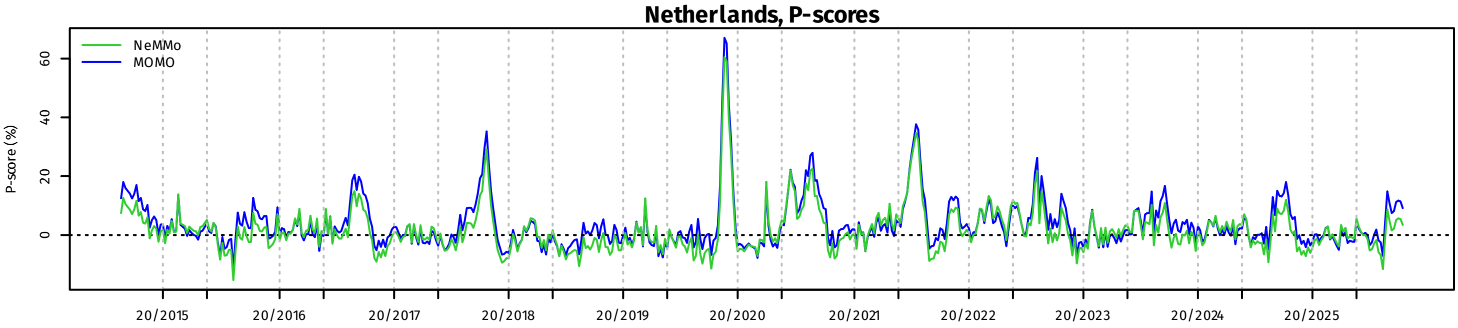
Netherlands, P-scores
NeMMo
MOMO
P-score (%)
0
20
40
60
20/2015
20/2016
20/2017
20/2018
20/2019
20/2020
20/2021
20/2022
20/2023
20/2024
20/2025

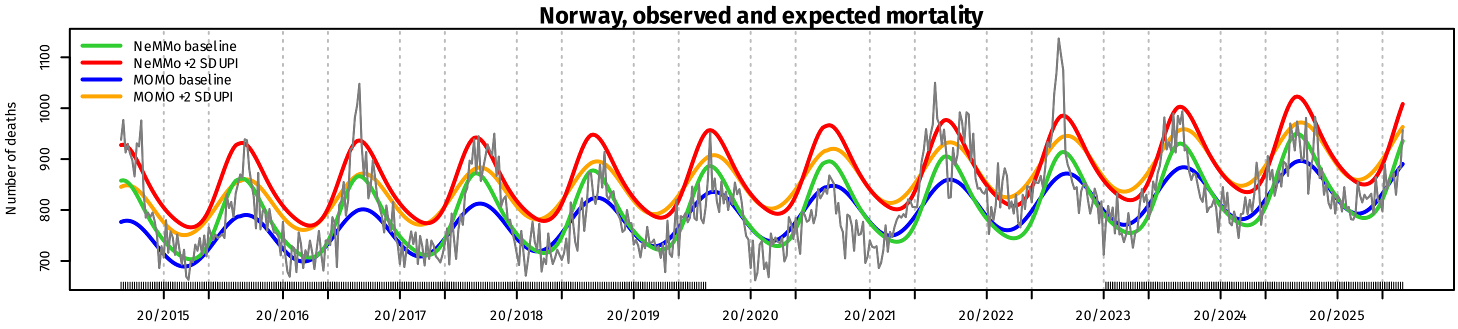
Norway, observed and expected mortality
NeMMo baseline
NeMMo +2 SD UPI
MOMO baseline
MOMO +2 SD UPI
Number of deaths
700
800
900
1000
1100
20/2015
20/2016
20/2017
20/2018
20/2019
20/2020
20/2021
20/2022
20/2023
20/2024
20/2025

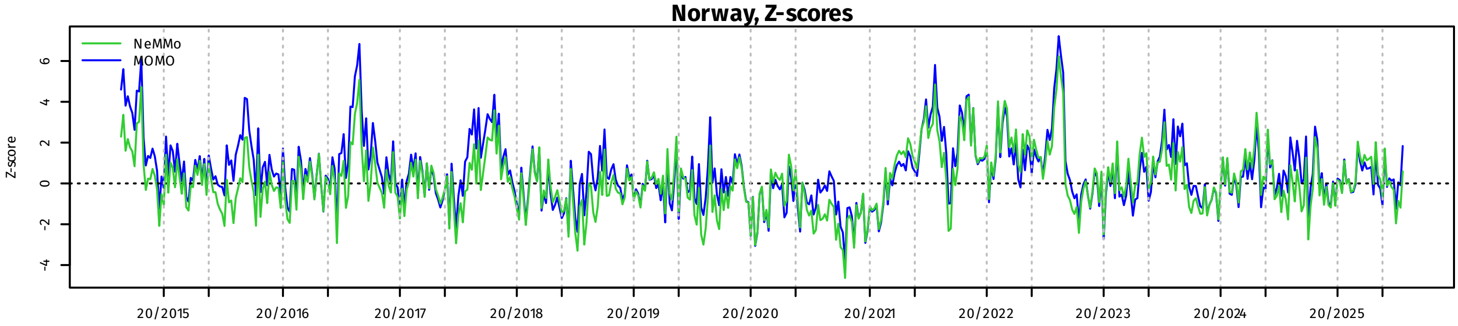
Norway, Z-scores
NeMMo
MOMO
Z-score
-4
-2
0
2
4
6
20/2015
20/2016
20/2017
20/2018
20/2019
20/2020
20/2021
20/2022
20/2023
20/2024
20/2025

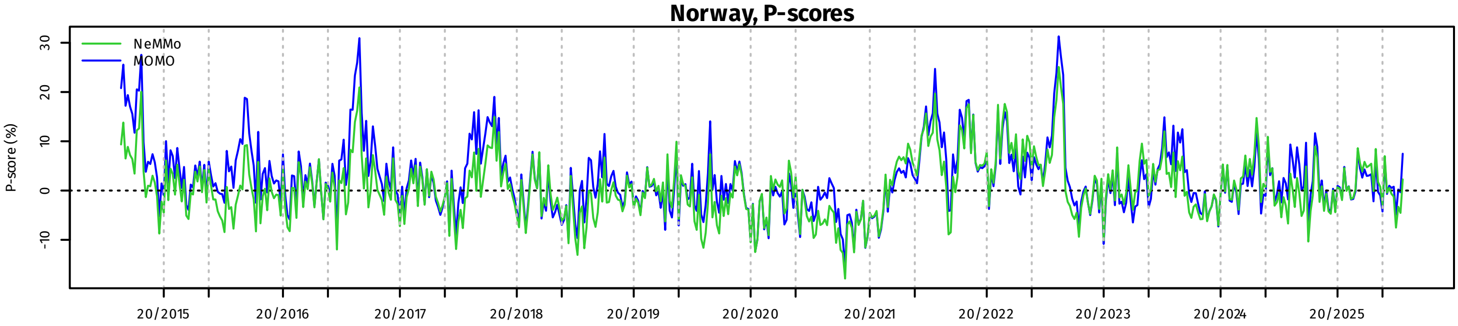
Norway, P-scores
NeMMo
MOMO
P-score (%)
-10
0
10
20
30
20/2015
20/2016
20/2017
20/2018
20/2019
20/2020
20/2021
20/2022
20/2023
20/2024
20/2025

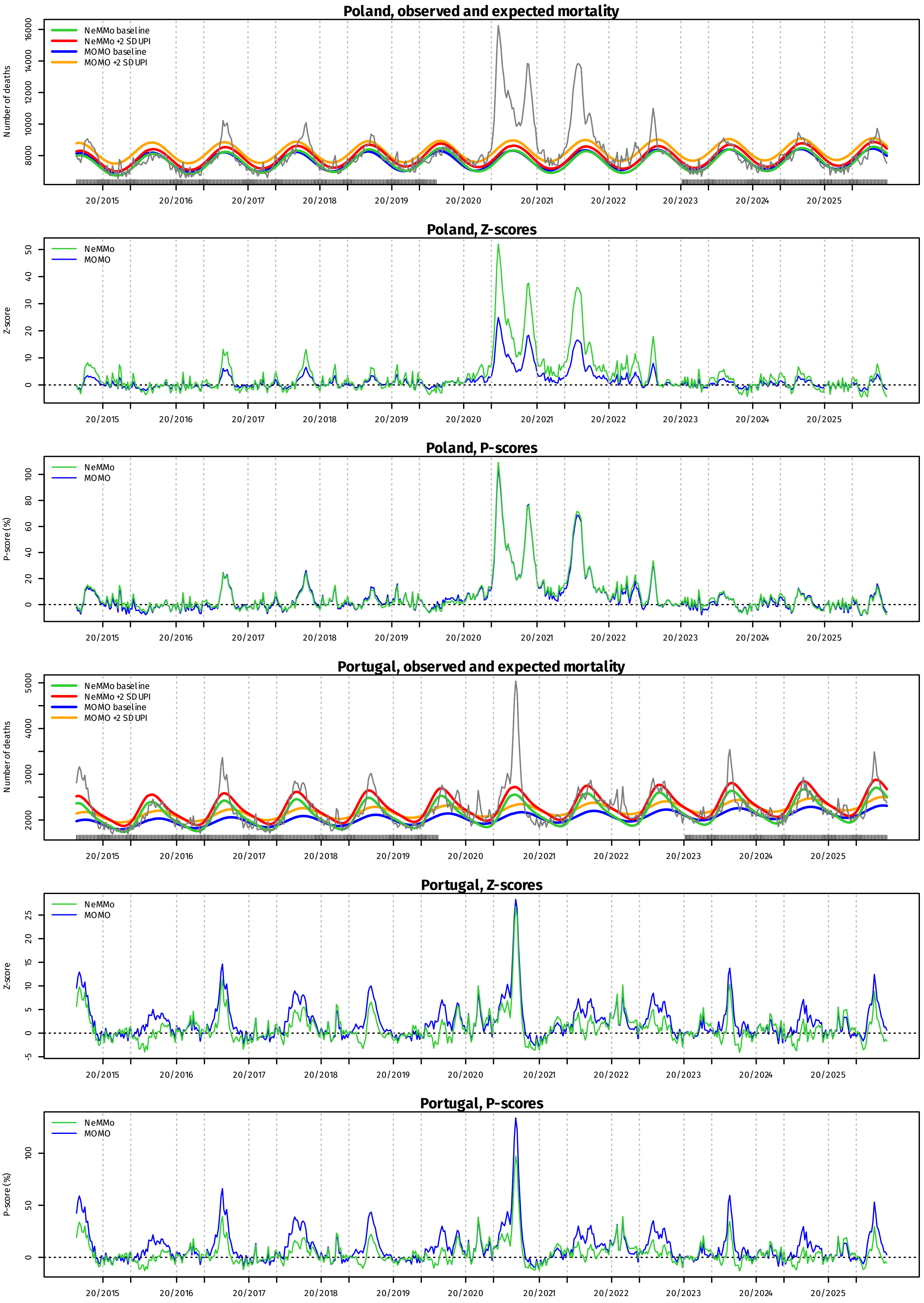
Poland, observed and expected mortality
NeMMo baseline
NeMMo +2 SD UPI
MOMO baseline
MOMO +2 SD UPI
Number of deaths
8000
10000
12000
14000
16000
20/2015
20/2016
20/2017
20/2018
20/2019
20/2020
20/2021
20/2022
20/2023
20/2024
20/2025
Poland, Z-scores
NeMMo
MOMO
Z-score
0
10
20
30
40
50
Poland, P-scores
NeMMo
MOMO
P-score (%)
0
20
40
60
80
100
Portugal, observed and expected mortality
NeMMo baseline
NeMMo +2 SD UPI
MOMO baseline
MOMO +2 SD UPI
Number of deaths
2000
3000
4000
5000
Portugal, Z-scores
NeMMo
MOMO
Z-score
-5
0
5
10
15
20
25
Portugal, P-scores
NeMMo
MOMO
P-score (%)
0
50
100

### Romania, observed and expected mortality

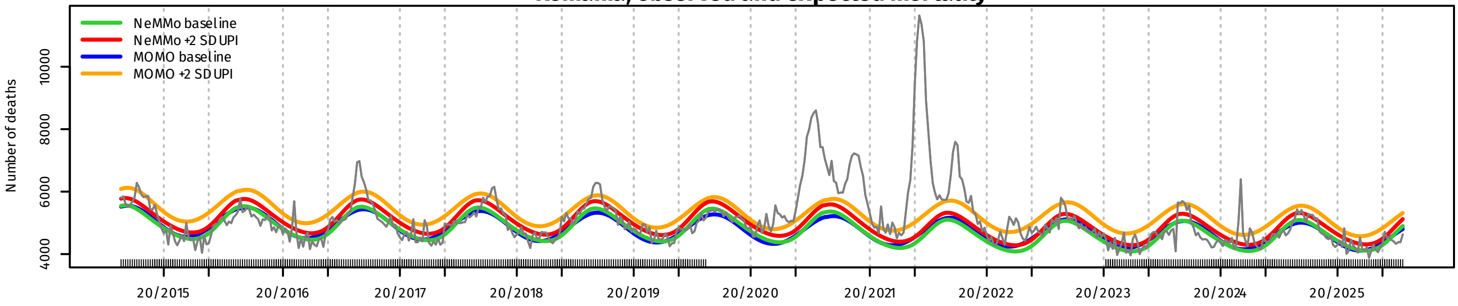


### Romania, Z-scores

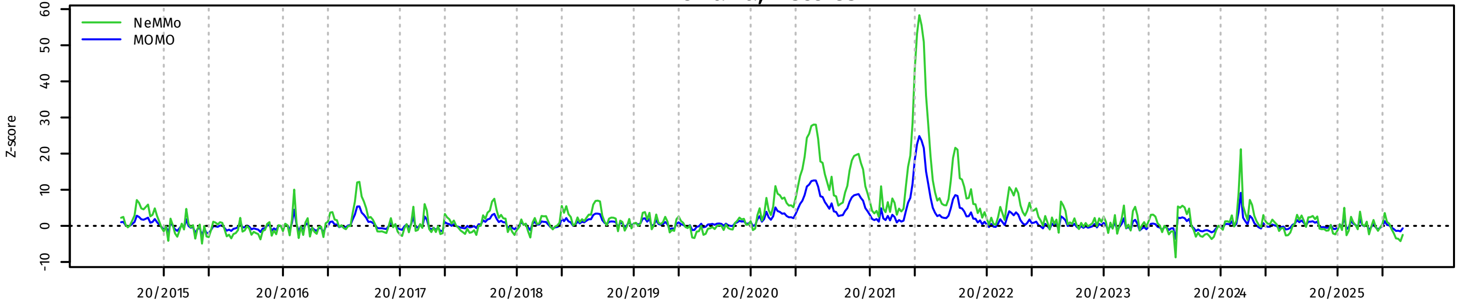


### Romania, P-scores

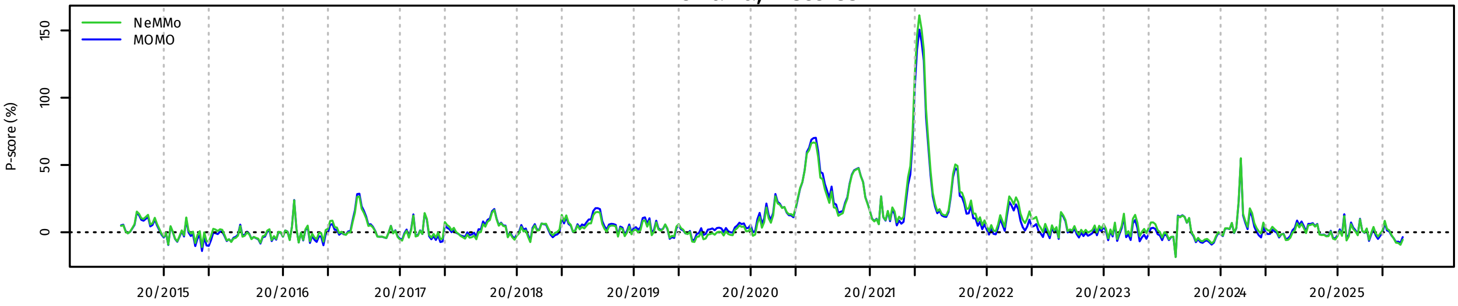


### Serbia, observed and expected mortality

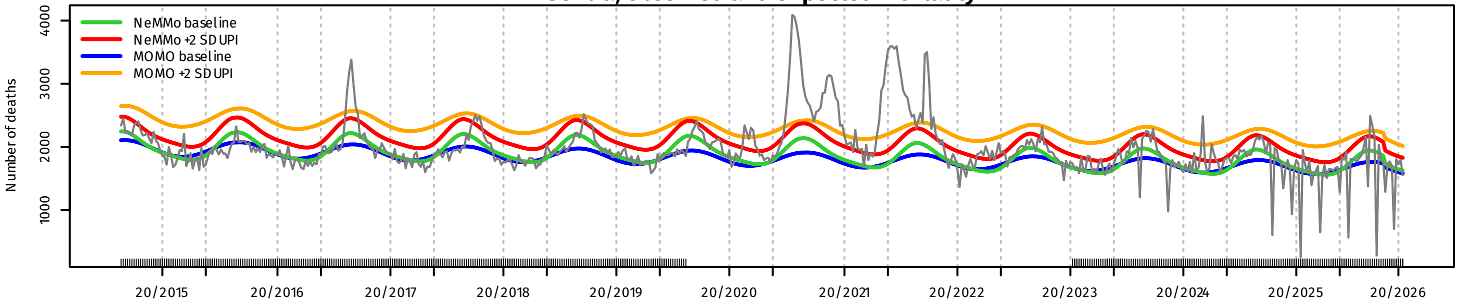


### Serbia, Z-scores

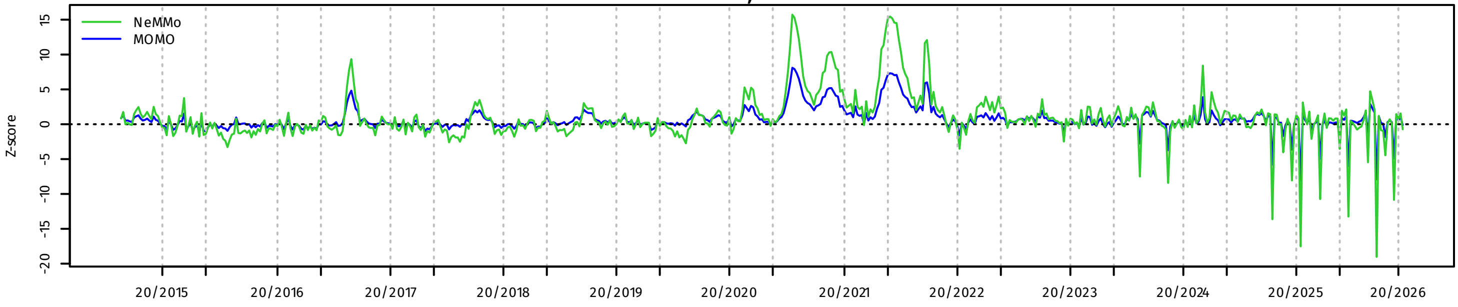


### Serbia, P-scores

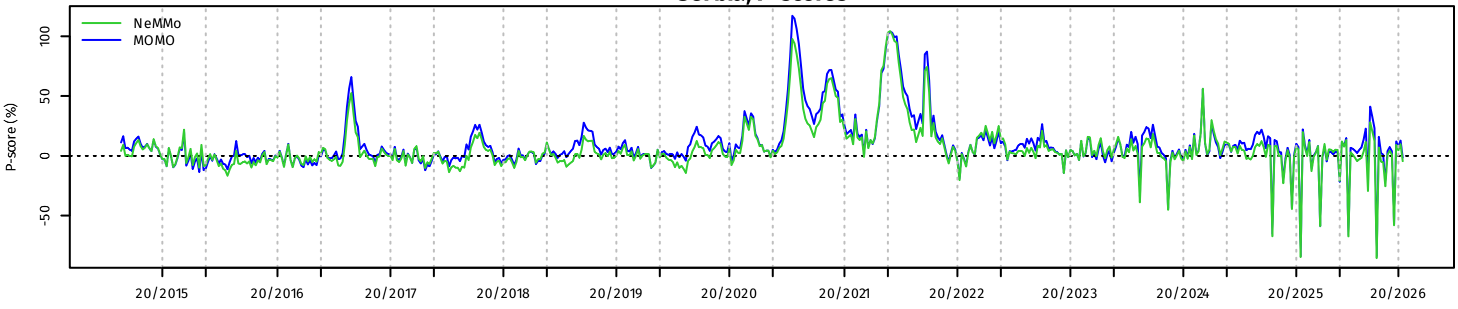

**Slovakia, observed and expected mortality**

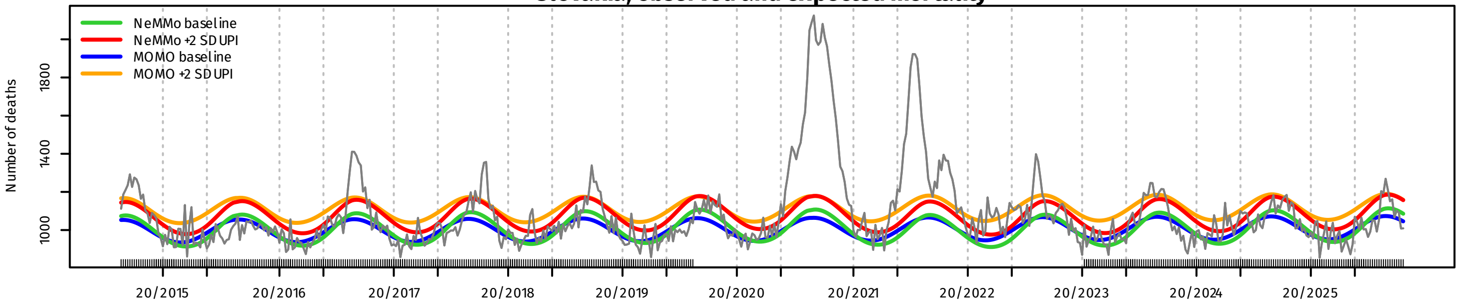


**Slovakia, Z-scores**

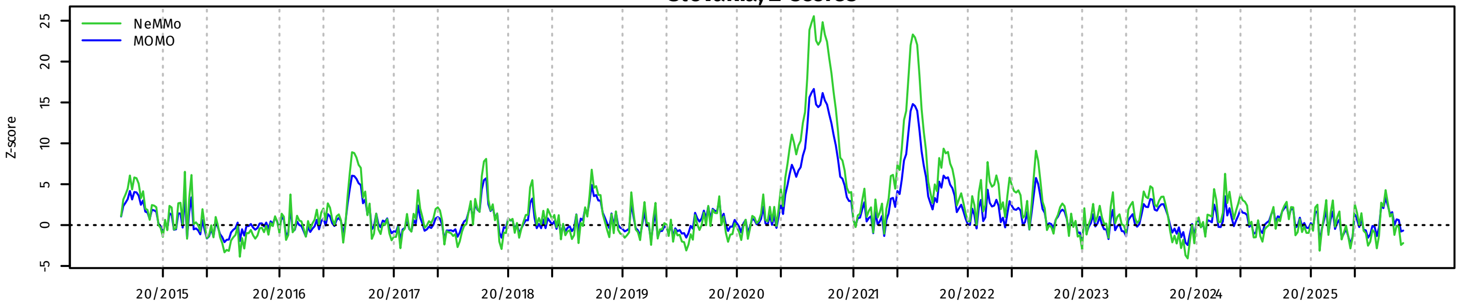


**Slovakia, P-scores**

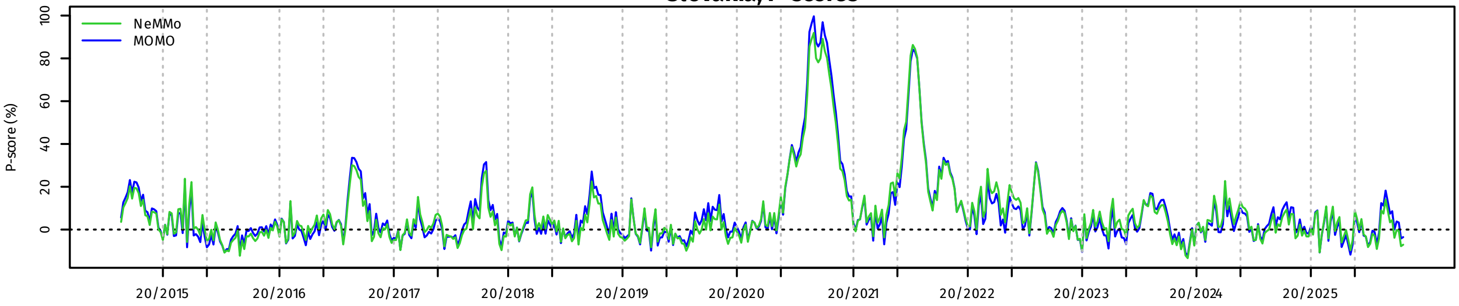


**Slovenia, observed and expected mortality**

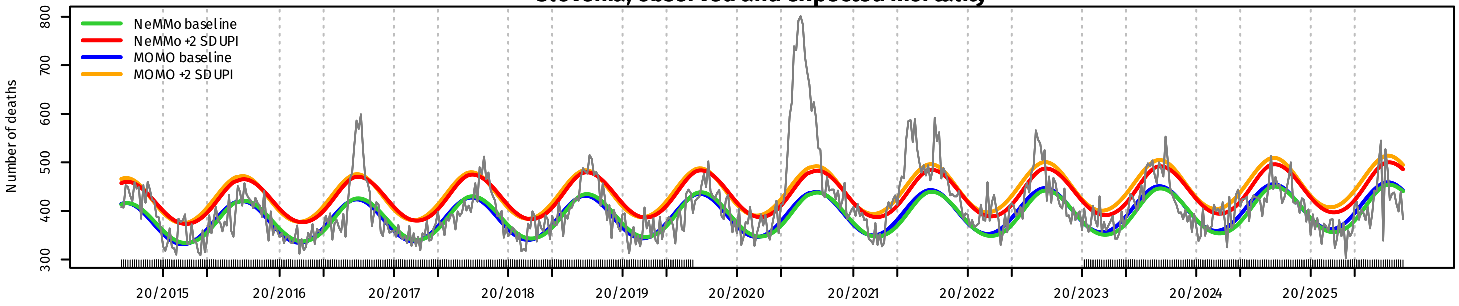


**Slovenia, Z-scores**

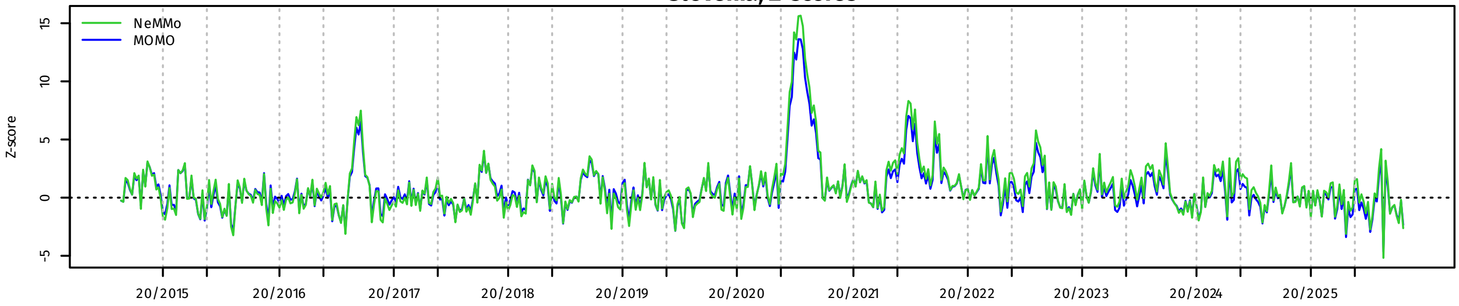


**Slovenia, P-scores**

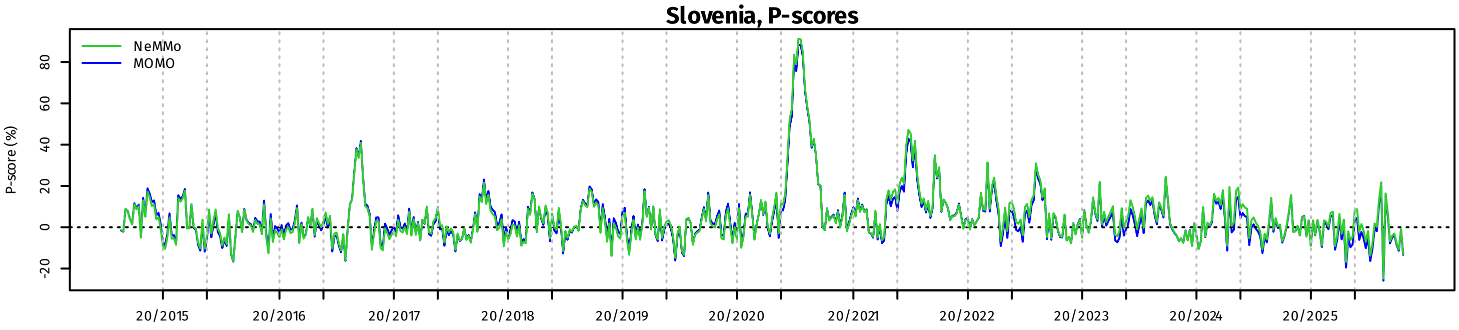

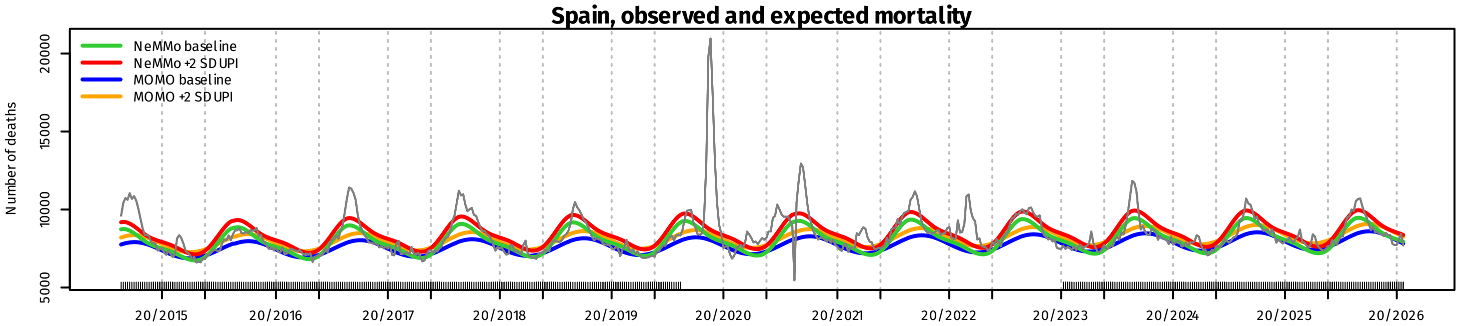
Spain, observed and expected mortality
NeMMo baseline
NeMMo +2 SD UPI
MOMO baseline
MOMO +2 SD UPI
Number of deaths
5000
10000
15000
20000
20/2015
20/2016
20/2017
20/2018
20/2019
20/2020
20/2021
20/2022
20/2023
20/2024
20/2025
20/2026

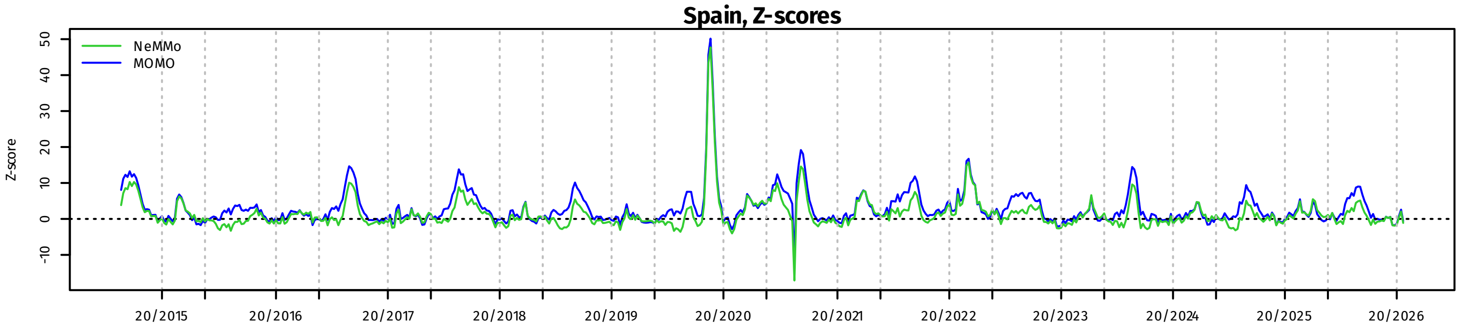
Spain, Z-scores
NeMMo
MOMO
Z-score
-10
0
10
20
30
40
50
20/2015
20/2016
20/2017
20/2018
20/2019
20/2020
20/2021
20/2022
20/2023
20/2024
20/2025
20/2026

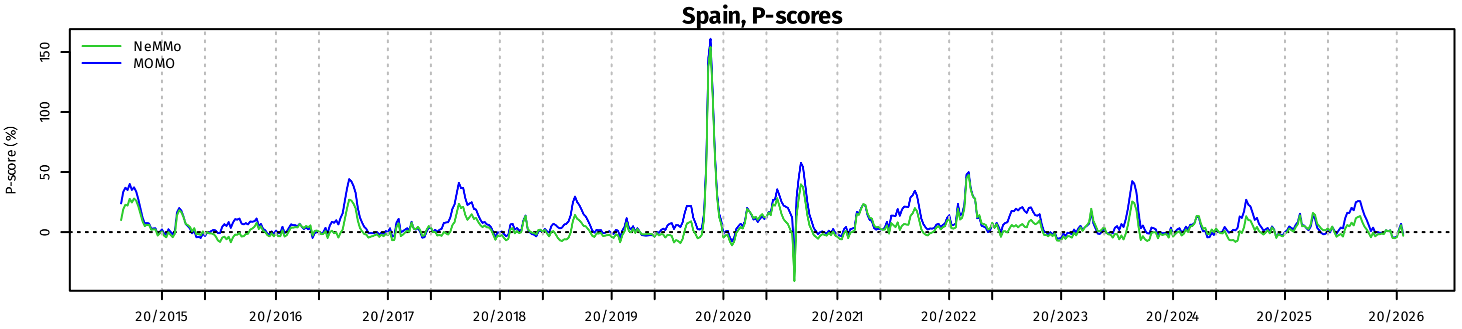
Spain, P-scores
NeMMo
MOMO
P-score (%)
0
50
100
150
20/2015
20/2016
20/2017
20/2018
20/2019
20/2020
20/2021
20/2022
20/2023
20/2024
20/2025
20/2026

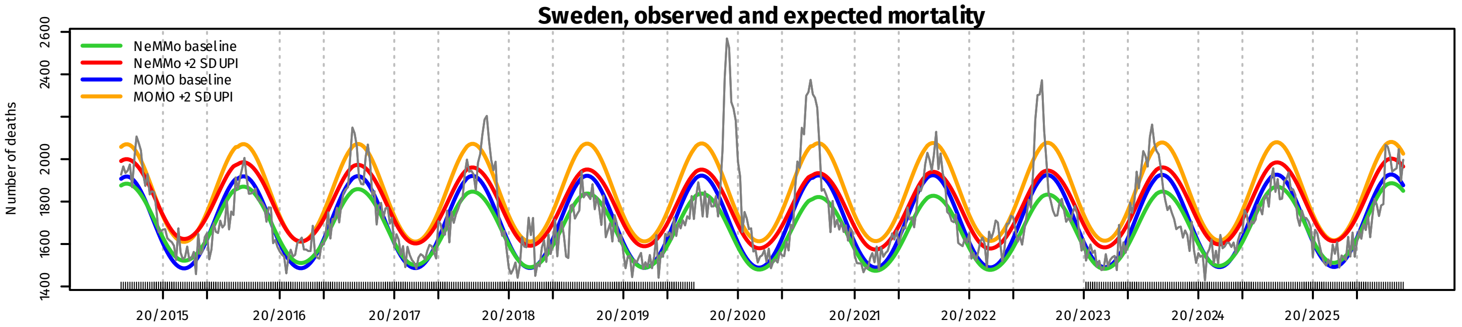
Sweden, observed and expected mortality
NeMMo baseline
NeMMo +2 SD UPI
MOMO baseline
MOMO +2 SD UPI
Number of deaths
1400
1600
1800
2000
2400
2600
20/2015
20/2016
20/2017
20/2018
20/2019
20/2020
20/2021
20/2022
20/2023
20/2024
20/2025

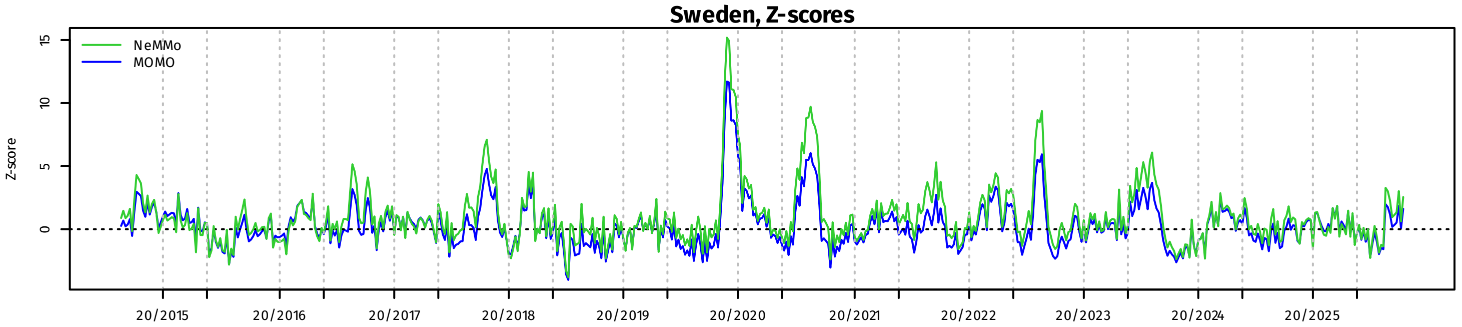
Sweden, Z-scores
NeMMo
MOMO
Z-score
0
5
10
15
20/2015
20/2016
20/2017
20/2018
20/2019
20/2020
20/2021
20/2022
20/2023
20/2024
20/2025

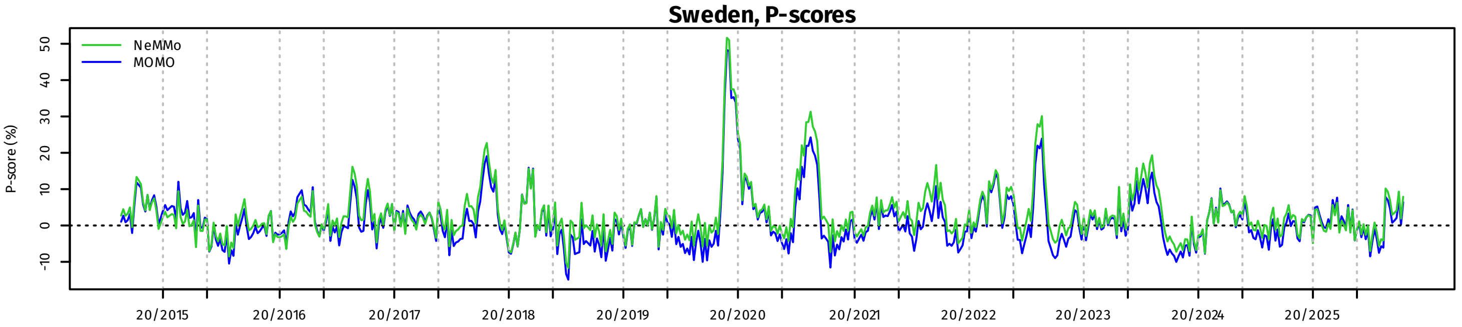
Sweden, P-scores
NeMMo
MOMO
P-score (%)
-10
0
10
20
30
40
50
20/2015
20/2016
20/2017
20/2018
20/2019
20/2020
20/2021
20/2022
20/2023
20/2024
20/2025

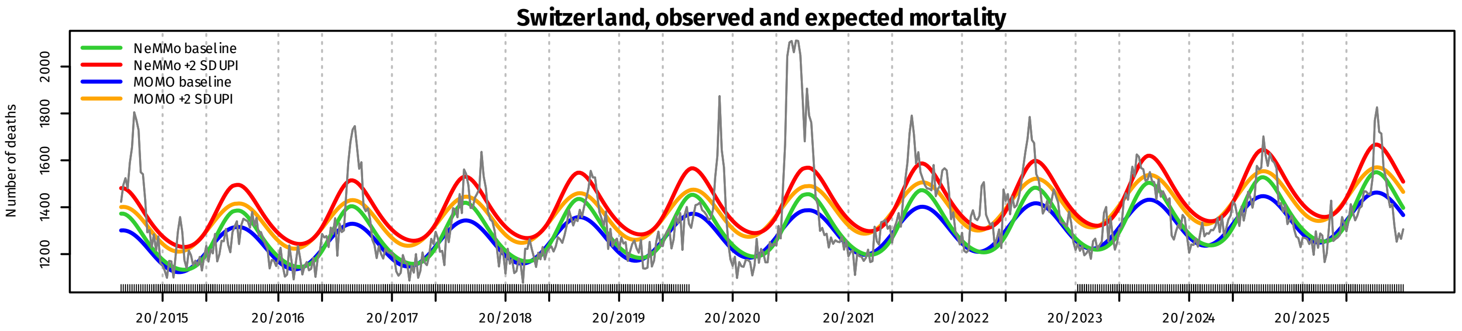
Switzerland, observed and expected mortality
NeMMo baseline
NeMMo +2 SD UPI
MOMO baseline
MOMO +2 SD UPI
Number of deaths
1200
1400
1600
1800
2000
20/2015
20/2016
20/2017
20/2018
20/2019
20/2020
20/2021
20/2022
20/2023
20/2024
20/2025

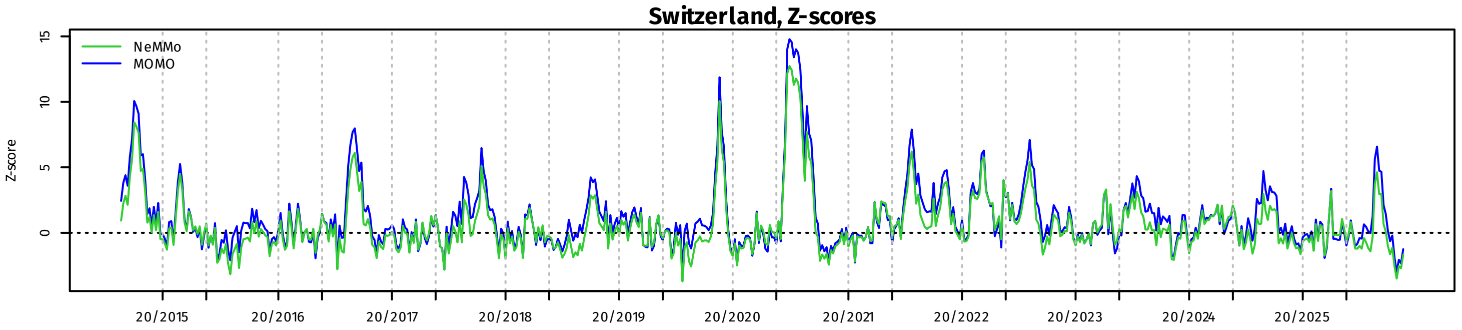
Switzerland, Z-scores
NeMMo
MOMO
Z-score
0
5
10
15
20/2015
20/2016
20/2017
20/2018
20/2019
20/2020
20/2021
20/2022
20/2023
20/2024
20/2025

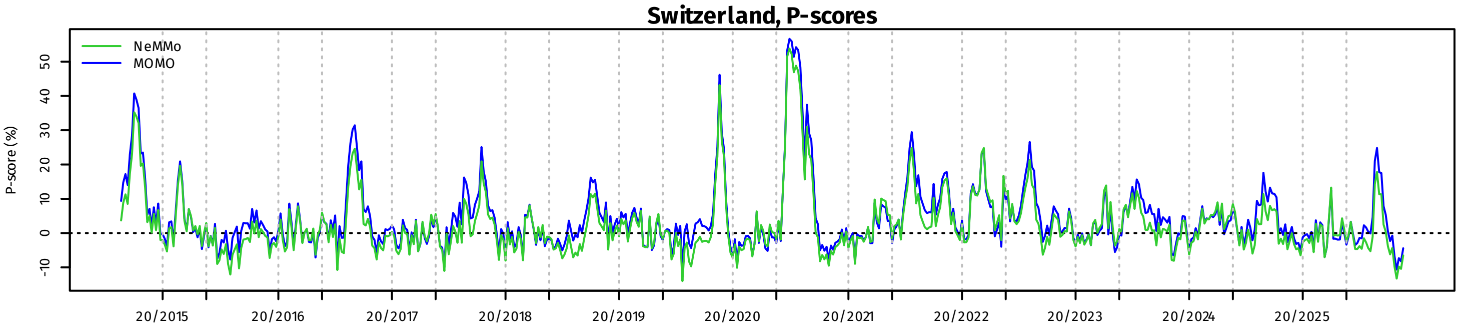
Switzerland, P-scores
NeMMo
MOMO
P-score (%)
-10
0
10
20
30
40
50
20/2015
20/2016
20/2017
20/2018
20/2019
20/2020
20/2021
20/2022
20/2023
20/2024
20/2025
