## Supplementary figures and images for "NeMMo: an improved statistical algorithm for excess all-cause mortality surveillance and monitoring"

### Supplementary Figure 2

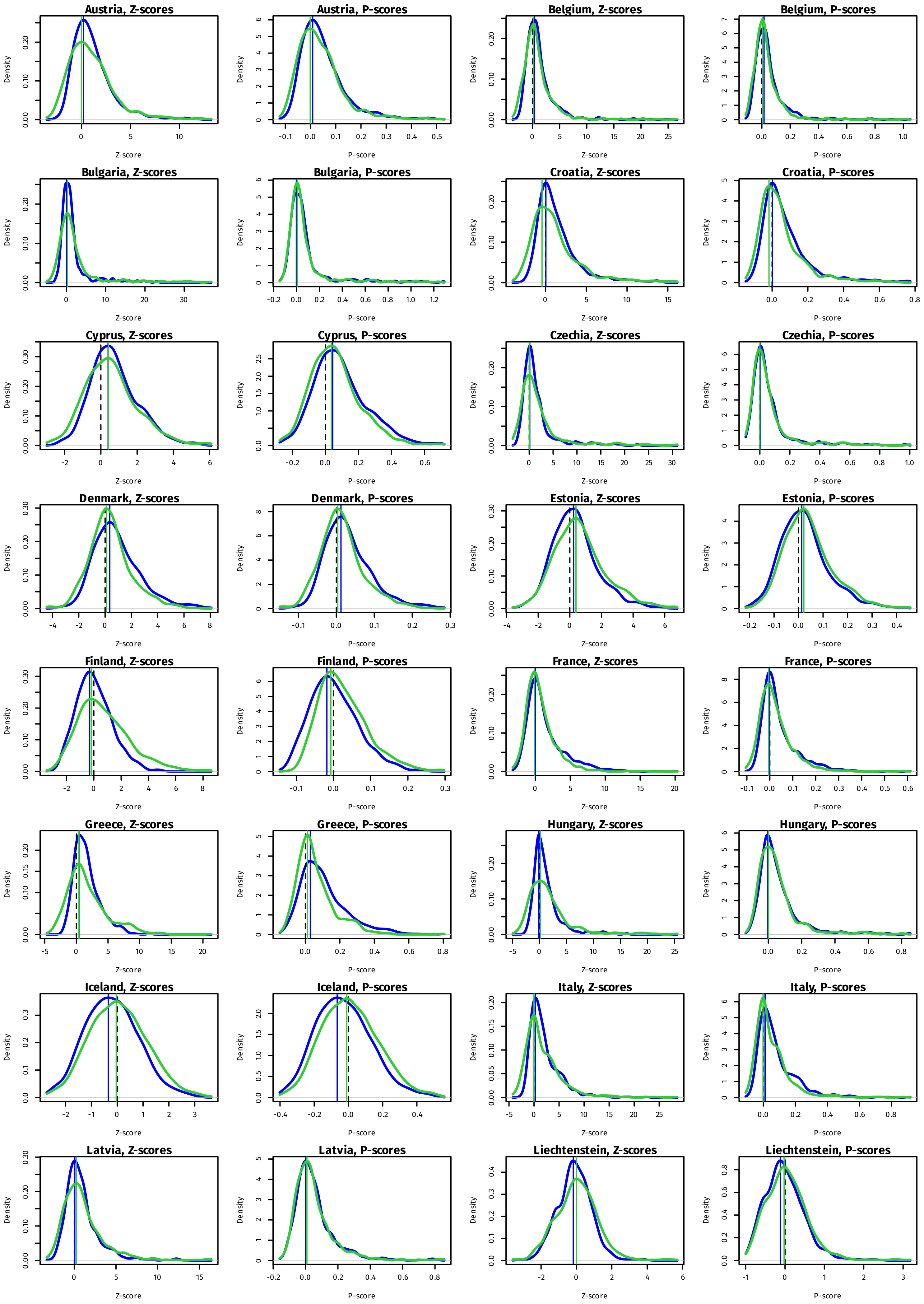

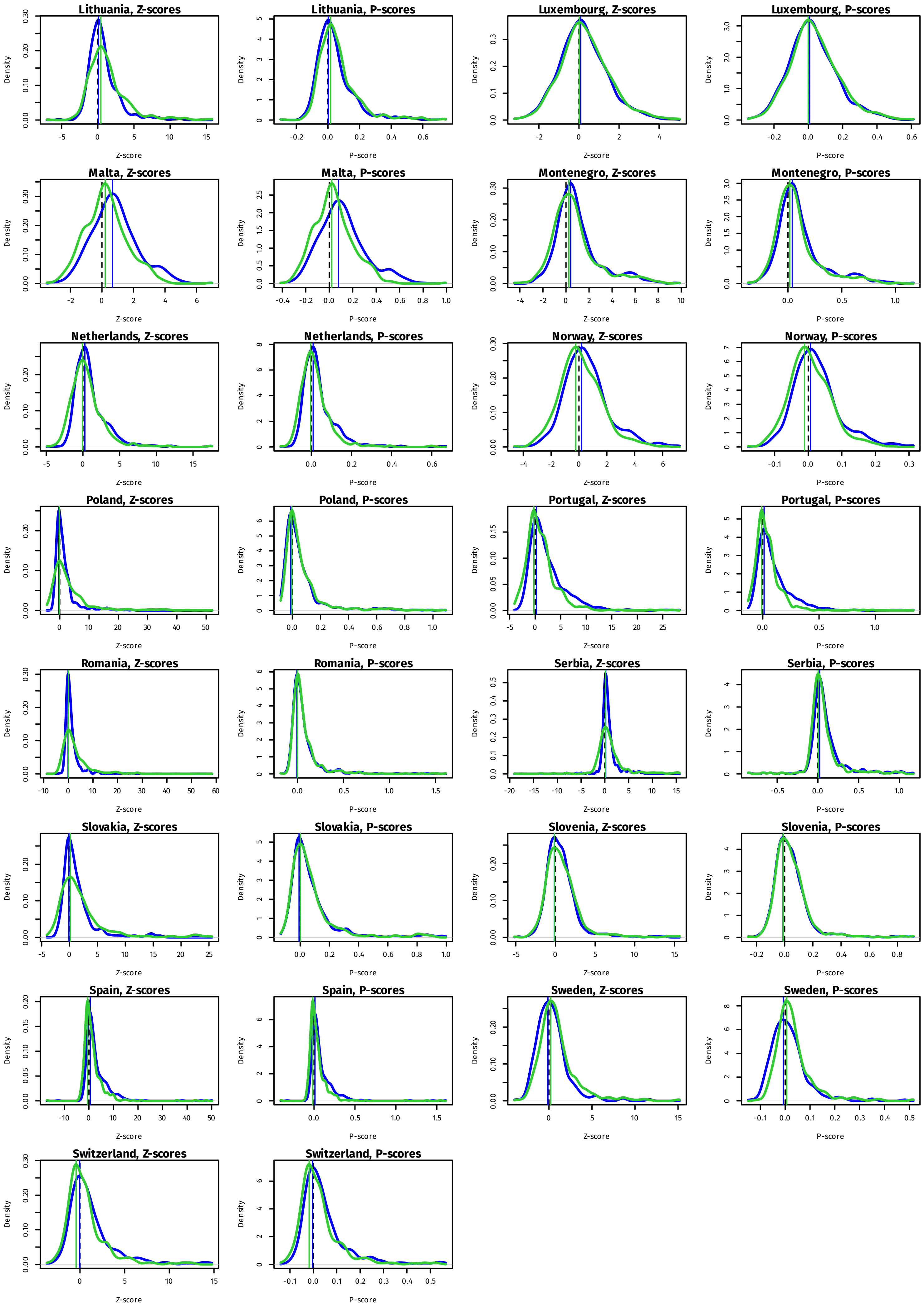
